# Population-scale patient safety data reveal inequalities in adverse events before and during COVID-19 pandemic

**DOI:** 10.1101/2021.01.17.21249988

**Authors:** Xiang Zhang, Marissa Sumathipala, Marinka Zitnik

## Abstract

Adverse patient safety events were associated with 110 thousand deaths in the U.S. alone in 2019. The COVID-19 pandemic has further challenged the ability of healthcare systems to ensure safe medication use, and its effects on patient safety remain unknown. Here, we investigate negative outcomes associated with medication use before and during the pandemic. Using a dataset of 10,443,476 reports involving 3,624 drugs and 19,193 adverse events, we develop an algorithmic approach to analyze the pandemic’s impact on the incidence of drug safety events by evaluating disproportional reporting relative to the pre-pandemic time, quantifying unexpected trends in clinical outcomes, and adjusting for drug interference. Among 64 adverse events identified by our analyses, we find 54 have increased incidence rates during the pandemic, even though adverse event reporting decreased by 4.4% overall. We find clinically relevant differences in drug safety outcomes between demographic groups. Compared to male patients, women report 47.0% more distinct adverse events whose occurrence significantly increased during the pandemic relative to pre-pandemic levels. Out of 53 adverse events with a pre-pandemic gender gap, 33 have an increased gender gap during the pandemic. While musculoskeletal and metabolic side effects are disproportionately enriched in women during the pandemic, immune-related adverse events are enriched only in men. We also find the number of adverse events with an increased reporting ratio is higher in adults (by 16.8%) than in older patients (adjusted for population size). Our findings have implications for safe medication use and tie the variation in adverse events to patients that may be disproportionately affected by preventable inequities during a public health emergency.

## Introduction

Adverse events from medications [1–10] accounted for over 110,000 deaths in the U.S. alone in 2019. Despite urgent implications [11–13], it remains largely unknown how the COVID-19 pandemic [14–22] influenced patient safety and what inequalities in diverse patient populations are exacerbated more than expected had the pandemic not occurred. Further, intricate dependencies between the pandemic’s effects, drugs, and patient characteristics [23–25] present a unique challenge for understanding patient safety during a public health emergency. Addressing this challenge can inform drug prescription, improve patient safety by identifying individuals at high risk for adverse events, and enable comparison of COVID-19 pandemic to other health emergencies to unveil the disruptive nature of public health crises and inform health policy. To this end, algorithmic approaches are needed to unveil how patient safety has changed with the pandemic onset and to compare patient safety to its pre-pandemic levels across patient groups and the entire range of human diseases and approved drugs.

Prior studies on adverse events have focused on laboratory environments, molecular characterization of drugs and target proteins, and limited clinical trial observations [26–29]. Patient safety studies during this pandemic are limited to very restricted pairs of drugs and adverse reactions, small numbers of reports, and narrow time ranges [30–32]. Further, such narrowly focused analyses can be confounded by historical biases in adverse event reporting and by mixing population groups that differ in their relative risks for clinical events.

We develop an algorithmic approach to systematically investigate negative outcomes associated with medication use and how they changed during the pandemic. Using a patient safety dataset of 10,443,476 adverse drug event reports spanning 7 years (Jan. 2013-Sept. 2020) and involving 3,624 drugs and 19,193 adverse events, our approach uncovers previously unknown impacts of the pandemic on patient safety and identifies variation of adverse events across patient groups. Disentangling confounders (such as sampling variance and temporal biases in the reporting of adverse events), our algorithmic approach detects gender- and age-related variations in adverse events and identifies demographic groups who are at higher or lower risk for adverse events during the pandemic than in the pre-pandemic time. This algorithmic effort leads to several key findings. We find substantial variation in adverse drug events before and during the pandemic. Among 64 adverse events identified by our analyses, we find 54 have increased incidence rates during the pandemic, even though adverse event reporting decreased by 4.4% overall. Further, we find that pre-pandemic gender differences are exaggerated during the pandemic. Women suffer from more adverse events than men relative to pre-pandemic levels, across all age cohorts. We also find relevant clinical differences in adverse event outcomes across age groups. For example, side effects, such as anxiety and insomnia, were disproportionately increased in women and the elderly, indicating they constitute at risk patient groups. Taken together, these analyses unveil risk-altering adverse effects that can inform drug prescription and public health policy, and enable comparison of this pandemic to other health emergencies. Finally, we present a comprehensive catalog of adverse events and their associations. The new resource can help discover relationships between drugs and safety events, especially in cases of rare events and effects within population subgroups that differ in their risks of specific clinical outcomes and are disproportionately affected by preventable inequities.

## Results

### Overview of our algorithmic approach for detecting differential patterns of drug response

We investigate 10,443,476 adverse event reports (involving 19,193 adverse events and 3,624 drugs) from the U.S. Food and Drug Administration (FDA) Adverse Event Reporting System (FAERS) dataset, collected from January 2013 to September 2020. To support safety surveillance of therapeutic products, the FAERS stores anonymized, manually reviewed adverse event reports received by the FDA. We use the dataset to detect adverse events that are significantly associated with the pandemic, pinpoint clinically relevant drugs strongly connected with adverse drug events, and identify disparities in the distribution of adverse events across sex and age. To this end, we develop an approach that identifies clinically meaningful adverse events that meet three criteria: i) the reporting frequency of the adverse event changed significantly between 2019 and 2020, ii) the change cannot be explained by its trend in previous years (2013 to 2019), and iii) the adverse drug reaction is strongly associated with at least one drug and cannot be explained by drug interference.

Corresponding to these three criteria, our approach contains three key components (Figure 1a). First, we estimate the reporting odds ratio of adverse events to detect those whose incidence has considerably shifted during the pandemic (Figure 1b; Eq. 2; Methods). Among those adverse events we then focus on those whose change in reporting frequency can not be explained by the expected upward or downward reporting trajectories had the pandemic not occurred. For that, we define Pandemic-Adverse Event Association Index (PAEAI) to quantify the effects of the pandemic on the incidence of adverse events (Eq. 3; Methods). We retain only adverse event associations with positive PAEAI values (Figure 1c), meaning that the their incidence in 2020 is significantly different than predicted on the basis of temporal trends. Third, our approach identifies adverse events with considerable associations to specific medications (Figure 1d; Eq. 4; Methods), omitting drugs withdrawn from the market in 2020. Resulting drug-adverse event pairs represent co-occurring drugs and adverse events whose reporting frequency has considerably changed during the pandemic (Eq. 5; Methods).

**Figure 1:**
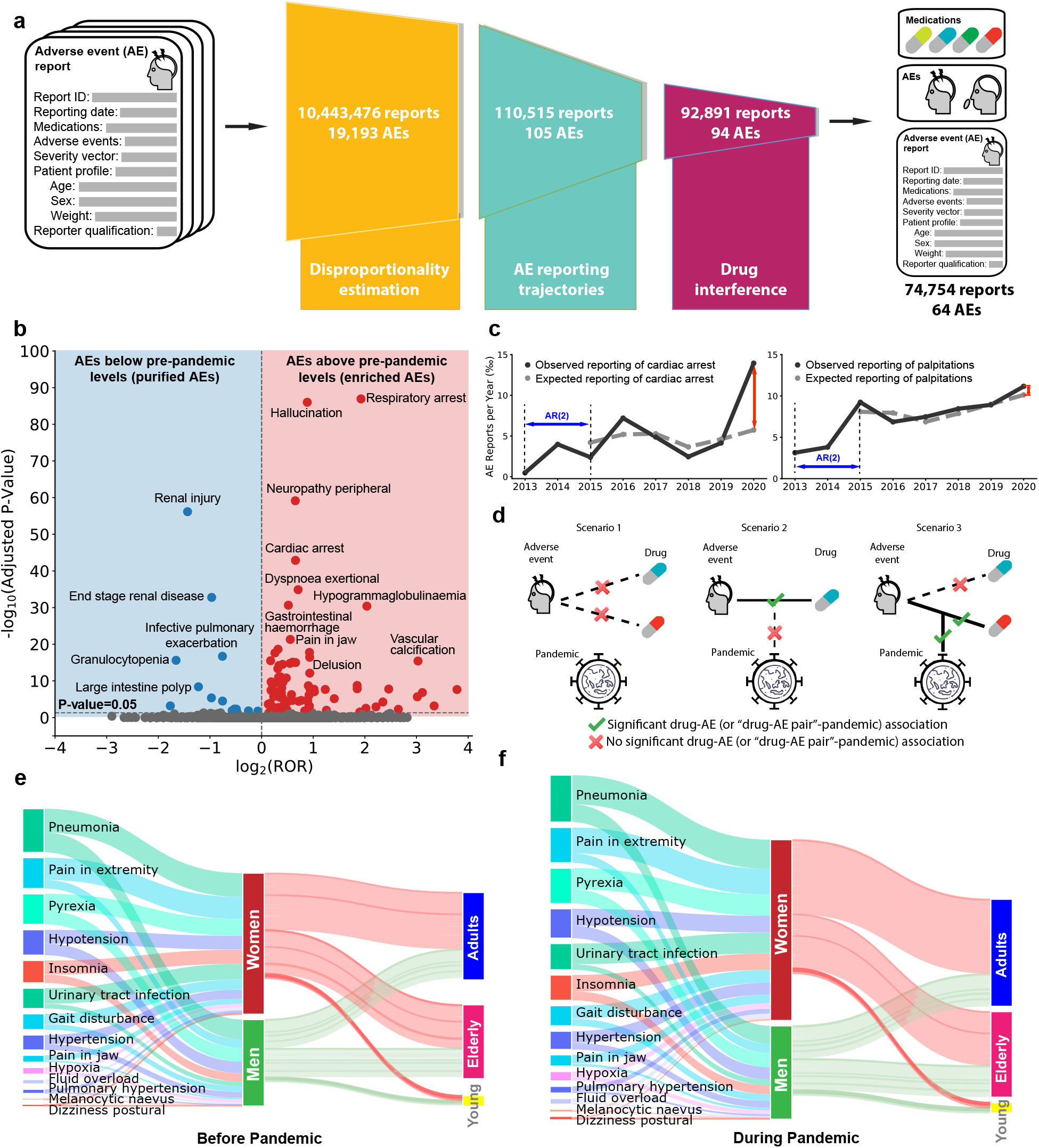
Population-scale model of patient drug safety. **(a)** Our algorithmic approach detects drug safety signals that are significantly associated with the pandemic by leveraging large-scale adverse event (AE) reports’ information about drugs and associated adverse reactions. The approach can be applied to any patient cohort; in the overall patient population, it identifies 64 among 19,193 adverse events. **(b)** Disproportionality estimation. Adverse events with p-value < 0.05 (Bonferroni-corrected) and whose 95% CI of ROR does not cross one are retained. In the overall population, this yields 105 adverse events: 72 enriched (overrepresented) and 33 purified (underrepresented). **(c)** AE reporting trajectories. We define PAEAI to characterize the temporal trend of adverse events’ reporting frequency (Methods) and only keep those with positive PAEAI, indicating a large margin between expected and observed reporting. Shown are trajectories of cardiac arrest (left; PAEAI =1.05; *R*^2^=0.49; keep) and palpitations (right; PAEAI = −0.54; *R*^2^=0.81; drop). **(d)** Drug interference. We first examine the association between adverse events and drugs, if significant, then measure the association between the formed drug-adverse event pair and the pandemic. We only retain adverse events that are significantly associated with at least one drug that makes the established pair significantly related to the pandemic (scenario 3). **(e)-(f)** Demographic information before and during the pandemic. The width of bars is proportional to the number of AE reports. Shown are adverse drug reactions with PAEAI > 0.8 and are enriched in women. The total number of reports in women significantly increased (p-value < 0.05, Student’s t-test) during the pandemic, but remained largely unchanged in men, implying an increased gender gap. The difference between widths of input and output streams in women/men are due to reports with unknown age. ROR: reporting odds ratio; CI: confidence interval; AR(2): second-order autoregressive model (see Methods).

Taken together, these three components constitute an algorithmic approach that can be used to analyze any patient cohort formed in a population, extract associations between drugs and adverse events, and quantify differential reporting patterns to identify high-risk demographics groups.

### Variation in adverse drug events before and during the pandemic

We find that most adverse events identified by our approach have increased reporting frequency during the pandemic, despite a 4.4% decrease in the total number of reports submitted by healthcare professionals from 220,920 in 2019 to 211,152 reports in 2020. Confirming the validity of our approach, the model detects a significant increase in the incidence of five adverse events directly related to COVID-19, such as coronavirus infection, coronavirus test positive, COVID-19, suspected COVID-19, and COVID-19 pneumonia (all p-value < 10^−37^, Fisher’s exact test): we exclude these from the rest of our analyses. The model detected 64 unique adverse events whose incidence changed during the pandemic in the overall population: 54 have increased reporting frequency during the pandemic, while only ten decreased in frequency (SI Figure S1). We define adverse drug reactions whose reporting frequency has disproportionately increased with the pandemic onset as enriched and those with significantly decreased reporting frequency as purified. Among the 54 enriched adverse events, delusion has the most significant association with the pandemic, with the largest PAEAI score of 1.95 (statistics for more adverse events in SI Table S1). The volume of FAERS reports involving drug-related premature delivery increased 73.4%, despite studies from the US and Europe finding a decrease in the overall incidence of preterm births during the pandemic [33–35]. Similarly, the incidence of drug-related bladder cancers increased by 147%, despite an overall decrease in cancer diagnoses after the onset of the pandemic [15]. The frequency of hallucination side effects increased 138% during the pandemic compared to before the pandemic, which could be related to reports linking paranoia about COVID-19 to hallucinations, as well as the neurological impacts of the disease itself [36, 37]. Our approach also detects large increases for severe side effects, such as respiratory failure and cardiac arrest. The domination of adverse events with increased frequency is consistently observed in most demographics across sex and age. For instance, in patients over 65 years old, 18 out of 19 identified adverse events are enriched, and only one is purified (SI Figure S1).

### Women suffer from more adverse drug events than men relative to pre-pandemic levels

While our approach detected 38 adverse events that increased in frequency in women (SI Table S3), only 16 enriched adverse events were detected in men (SI Table S2; Figure 2a). This finding is consistent with that 62.0% of all reports in FAERS are from women and only 38.0% from men (excluding reports with unknown sex). Even after normalizing to the greater number of female patients in FAERS, there are still 48.5 enriched adverse events per million female patients, which is 47.0% higher than 33.0 per million male patients. Further, 32 of the 38 side effects are enriched only in women, while only 10 out of 16 are enriched only in men (Figure 3a). The model identifies six adverse events enriched in both women and men. For example, respiratory arrest has a similar PAEAI in males (0.80) and females (0.74), suggesting that pandemic has a comparable influence on men and women regarding respiratory arrest incidence. In contrast, the confusional state has a higher PAEAI in women (0.94) than in men (0.26), suggesting that changes brought on by the pandemic exert greater influence on the incidence in female patients.

**Figure 2:**
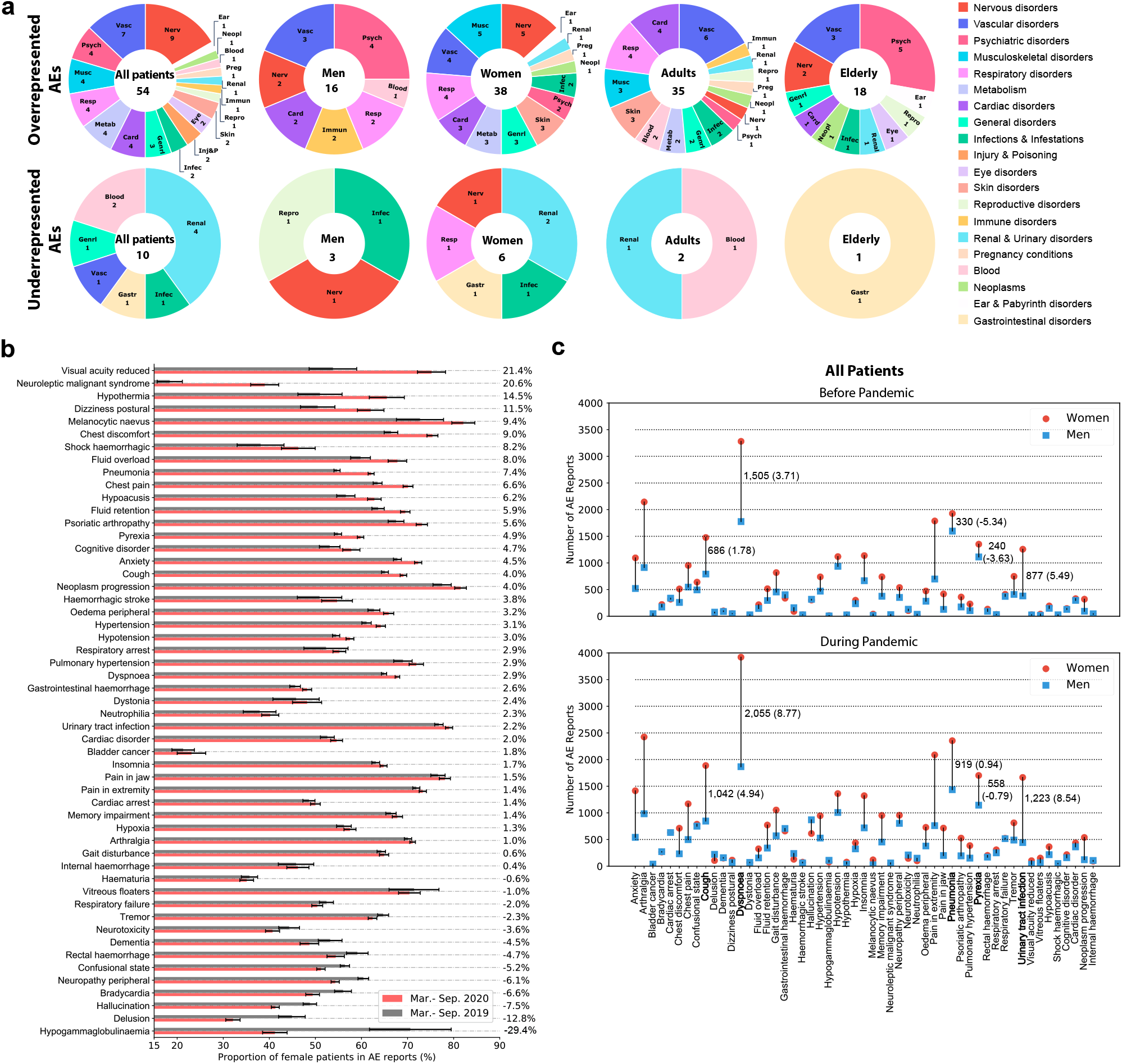
Distribution of detected adverse events. **(a)** Distribution of adverse drug reactions across System Organ Classes (SOC). Values listed indicate the number of averse events associated with each patient cohort and are divided into groups by etiology of adverse events (Methods). In the overall population, nervous and vascular SOCs are most common, with nine and seven side effects, respectively. The representation of enriched psychiatric adverse events varies: four out of 16 adverse events in men (25%) and five out of 18 in the elderly (27.8%), compared to only two out of 38 in women (5.3%) and one out of 35 in adults (2.9%). There is one overrepresented and one underrepresented drug side effect in young patients (SI Figure S2-S3). **(b)** The proportion of female patients in the 53 side effects enriched in the overall population (omitting reports with unknown sex and excluding premature delivery which only occurs in women). In the majority (75.5%) of side effects, female patients account for a higher proportion of reports during the pandemic, compared to before the pandemic. **(c)** Changes to gender disparities in the 53 side effects enriched in the overall population during the pandemic (same 53 as in panel b). The gap in the number of reports for men and women is exacerbated during the pandemic in most (41/53) of the adverse drug events. We annotate the top five adverse events that have the largest increase (bolded), with the first number indicating the absolute difference in number of reports, and the number in parentheses showing the difference normalized by population size. For example, a normalized gap of 4.08 in dysponea indicates there are 4.08 more reports per thousand women than per thousand men (negative numbers indicate the reporting frequency in women is smaller than in men, even though the absolute number of reports may be higher in women). Considering incidence proportion (the number of reports per thousand patients) of adverse events (SI Figure S4): we find that the gender disparity is enlarged in 33 out of 53 the adverse drug reactions during the pandemic, consistent with the trend observed in the absolute number of reports.

**Figure 3:**
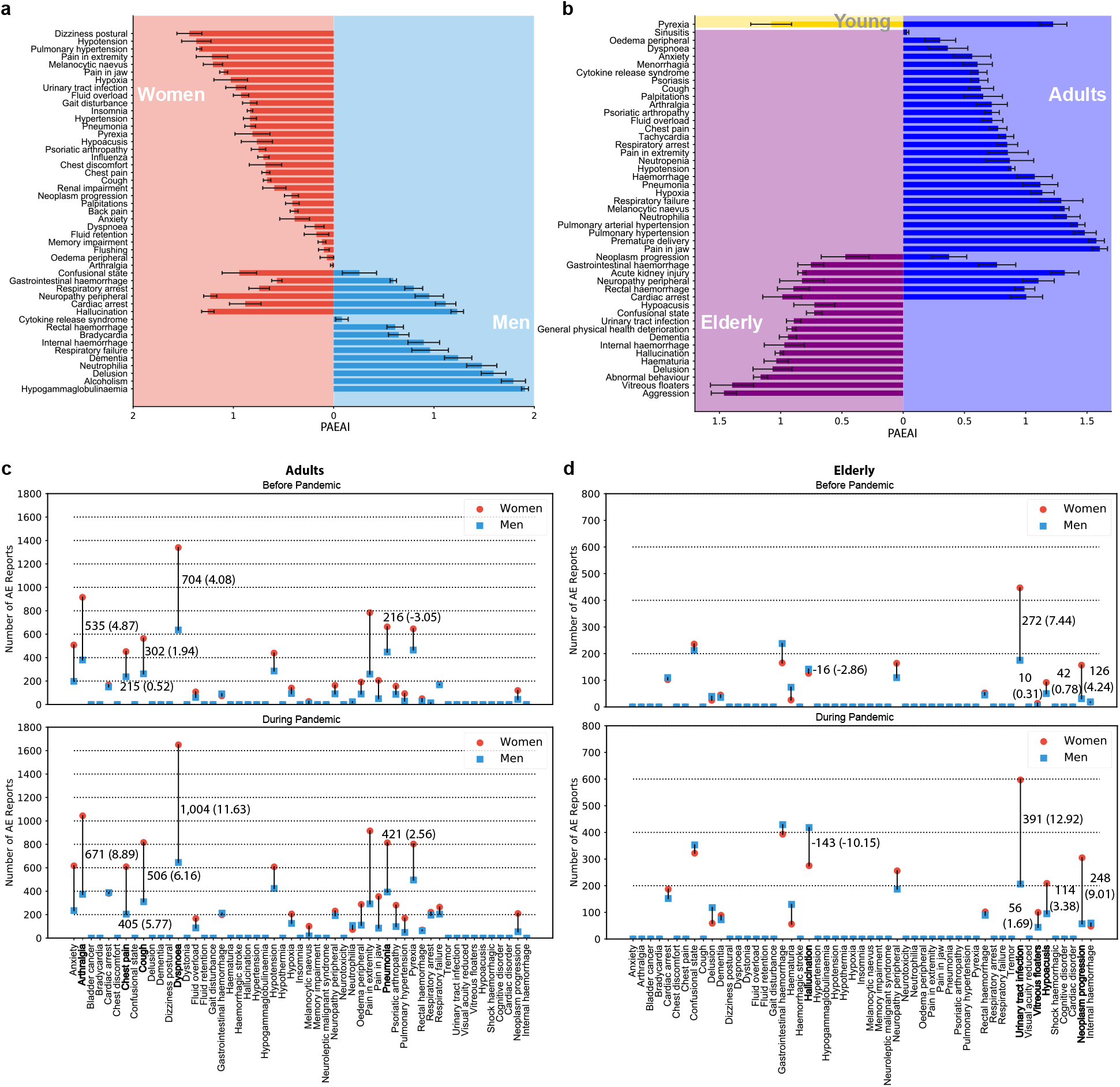
Disparities across gender and age. **(a)** PAEAI values differ in men and women. Higher PAEAI indicates the incidence of a drug-related side effect has undergone a greater change during pandemic than pre-pandemic and that change cannot be explained by temporal trends. We find 10 adverse drug reactions show significant association with the pandemic only in men but not in women, whereas 31 are associated with the pandemic only in women. **(b)** Age differences in PAEAI. We find that adults have more drug adverse events whose reporting frequency was impacted by the pandemic, compared to the young and elderly cohorts. Pyrexia is significantly associated with the pandemic in both young and adult cohorts. There are no side effects impacted by the pandemic in both young and elderly individuals. The algorithmic approach detects six adverse drug reactions impacted in both adults and the elderly. The majority (28/35, 80.0%) of adverse events observed in adult patients are not found in other age groups. In contrast, there are 12 out of 18 drug reactions in elders are not found in adults. **(c)** Gender gap in adults (omitting reports with unknown sex). We detected 35 enriched adverse events in adults, and omitted premature delivery and 10 that were only observed in reports with unknown sex: leaving 24 with gender difference. Pre-pandemic gender disparities are increased in 21 out of 24 (87.5%) adverse events, in response to the pandemic. Of these 21 adverse events, 18 have more reports involving women than men, suggesting a disproportionate vulnerability in adult women for gender disparities. Regarding incidence proportion, we note that the gender gap is magnified in 70.8% of the adverse events (SI Figure S5). **(d)** Gender gap in the elderly (omitting reports with unknown sex). In 13 out of 14 adverse drug events (18 adverse events significantly associated with the pandemic in elderly patients, 4 only observed in unknown sex and omitted), a pre-existing gender gap is intensified during the pandemic. Five among these drug side effects (5/13, 38.5%) are reported in more males than females during the pandemic, while only 3 out of 24 side effects (3/24, 12.5%) in adults: indicating the gender inequality is mitigated in aged people in contrast to adults. With the onset of pandemic, gender discrepancy (in terms of incidence proportion) has worsened in 11 out of 14 adverse events where gender inequality previously existed (SI Figure S6; adjusted for population size).

### Proportion of female patients increased across most enriched adverse events

Next, we assess the proportion of female patients in each of 54 side effects that were enriched in the overall population, comparing the proportion before and during the pandemic (Figure 2d). We exclude premature delivery, for which all patients are female. We find the proportion of female patients increased in 40 out of 53 adverse events during the pandemic. The two adverse events with the largest increases are reduced visual acuity, where the proportion of women increased from 53.8% to 75.2%, and neuroleptic malignant syndrome, where the proportion of women increased from 18.4% to 39.0%. We find the proportion of women in drug-related anxiety reports increased from 67.8% to 72.3%, confirming that women report anxiety at higher rates during the pandemic than men [38]. We observe an increase in the proportion of reports for side effects associated with anxiety disorders: insomnia, dyspnoea (shortness of breath), and dizziness, but find a decrease in side effects commonly associated with psychosis disorders: delusion, hallucinations, and dementia.

Our analyses identify 13 adverse events for which the proportion of female patients has decreased while male patients’ proportion increased (SI Figure S7). For example, the proportion of hypogammaglobulinemia reports with female patients dropped steeply from 70.6% to 41.2% during the pandemic. As patients with hypogammaglobulinemia are immunocompromised and at higher risk for COVID-19, this finding warrants investigation into why the incidence of hypogammaglobulinaemia decreased in women and if there is an increase in undiagnosed cases.

### Variation in adverse drug events across age groups

Stratification of patients by age groups (Figure 3b) reveals one enriched adverse event in young patients (SI Table S4), 35 in adults (SI Table S5), and 18 in elderly patients (SI Table S6). Even after accounting for the differences in each patient cohort’s size, there are still 68.8 side effects per million adults with increased reporting frequency during the pandemic, which is greater than the 27.8 side effects per million young patients and 58.9 adverse events per million elderly. The one side effect enriched in young patients, pyrexia (PAEAI =1.08), is similarly enriched during the pandemic in adults (PAEAI =1.22) but is not significantly impacted in the elderly.

Twenty-eight out of the 35 enriched adverse events in adults are unique to adults (not associated with young or elderly patients). For example, drug-related jaw pain has increased incidence in adult patients during the pandemic (PAEAI = 1.61) but not in young or elderly patients. Six adverse drug reactions are enriched in both adult and elderly patients (cardiac arrest, rectal hemorrhage, neuropathy peripheral, acute kidney injury, gastrointestinal hemorrhage, and neoplasm progression). While most have similar PAEAI scores in both cohorts, acute kidney injury has a PAEAI of 1.28 in adults in contrast to 0.81 in elderly patients, suggesting an age-related disparity in the pandemic’s impact on drug-related kidney injury. Twelve adverse events are uniquely enriched in the elderly, including five mental health-related events (hallucination, delusion, aggression, abnormal behavior, and dementia). This finding is in contrast with earlier surveys [39, 40] showing lower rates of overall anxiety-, depression-, and stress-related disorders in the elderly compared to younger age groups and suggests that drug-induced psychiatric effects may need to be addressed differently by healthcare systems.

### The impact of adverse drug reactions on human organs during the pandemic

We divide 54 adverse events found to be enriched in the overall population into 19 categories based on the System Organ Classification (SOC; [41]) (Figure 2a; SI Figure S2-S3). Nervous system and vascular side effects are the most common, with nine and seven adverse events, respectively, suggesting the incidence of these two adverse event classes are more susceptible to changes brought on by the pandemic. This finding points to the evidence suggesting COVID-19 has a significant impact on the vasculature and increases the risk of developing neurologic disorders [42, 43].

Ten adverse events have their reporting frequency disproportionately decreased during the pandemic, four of which are urinary system disorders and two are blood-related adverse events (SI Figure S3). Although the incidence proportion are decreased in reports submitted by professional healthcare workers, we find five adverse events (infective pulmonary exacerbation of cystic fibrosis, chronic kidney disease, osteonecrosis of jaw, renal injury, and nausea) have more self-reported cases (submitted by customer themselves) during the pandemic in contrast to before pandemic (SI Figure S8-S9). In particular, reports of kidney injury have dropped dramatically among professionals, but have risen sharply among non-professionals, suggesting an unmet need for pharmacovigilance in drug-related renal injury during the pandemic.

Next, we investigate the SOC classes of adverse events enriched in demographic groups. (Figure 2a). Our approach identifies 38 adverse events that are enriched in female patients and distributed across 14 SOC classes with most common being nervous system, musculoskeletal, vascular, and respiratory disorders (SI Figure S2-S3). In male patients, 16 enriched adverse events spread across seven SOC classes with most common being vascular and psychiatric disorders. Blood and immune system disorders are enriched only in men but not women. In contrast, side effects in nine SOC classes are only overrepresented in women, but not men, including metabolism, musculoskeletal, skin, infections, and pregnancy-related disorders. Although psychiatric adverse events are enriched in both men and women, there are four psychiatric side effects in men, and only two in women: hallucination is enriched in both; the incidence of anxiety is increased in females but not in males, while alcoholism, delusion, and dementia are only enriched in male patients.

Psychiatric adverse events are overrepresented in the elderly, with five enriched adverse events, compared to only one in adults: lending further support to our finding that the elderly may be differentially susceptible to psychiatric side effects of medications. In particular, hallucination, delusion, abnormal behaviour, aggression, and dementia are enriched in the elderly but not adults, while anxiety is only enriched in adults. Moreover, in the elderly, we observe one eye- and one ear-related adverse events (vitreous floaters and hypoacusis, respectively), but do not detect either in adults. Across sex and age cohorts as well as in the overall population, vascular-related side effects are enriched, suggesting it is a class of adverse drug reactions whose incidence is widely impacted by the pandemic.

### Pre-pandemic gender differences are exaggerated during the pandemic

We further investigate gender disparities among the enriched adverse events, and whether any pre-existing gender gaps changed during the pandemic. In the overall population, gender disparities are observed in all of the 53 adverse events before the pandemic, and we find that these gender disparities are exacerbated during the pandemic in 41 out of 53 adverse events (Figure 2c). For example, before the pandemic there were 330 more female patients report drug-related pneumonia than male patients: during the pandemic, the gap nearly tripled to 919 reports. The gender gaps normalized by population size are shown in SI Figure S4-S6. Similarly, there are 877 more reports from female patients of urinary tract infections (UTIs) than male, consistent with anatomical and clinical evidence that women are more susceptible to UTIs [44]; the gender gap has increased to 1,223 reports during the pandemic. In contrast, hallucination showed almost no gender disparity (15 cases) before the pandemic. Yet after the pandemic, male patients are overrepresented compared to women with a large margin (243 cases). Among the 41 adverse events with an increasing gender gap, 33 involve more female patients than male patients during the pandemic (Figure 2c), which is consistent with the historical exclusion of women from clinical trials [3]. During the pandemic, the largest gap is observed in dyspnoea, with 3,921 reports from female patients and only 1,866 reports from male patients. The second-largest gap is observed in arthralgia with 2,424 reports involving women and only 983 reports for men.

Considering adults and elderly, we observe a similar increase in pre-existing gender disparities. Among 35 adverse events enriched in adults during the pandemic, 24 have gender inequality (10 are only reported in unknown sex, excluding premature delivery). We find that 21 out of 24 adverse events have a larger gender gap after the onset of the pandemic. Among 21 drug reactions with exacerbated gender difference, 18 involve more women than men during the pandemic (Figure 3c; Figure S5 is adjusted for population size). In elderly patients (Figure 3d; Figure S6 is adjusted for population size), gender disparities exist in 14 out of 18 enriched adverse events (four are only observed in unknown sex). The gender disparities increased in 13 out of 14 drug reactions during the pandemic; eight are female-related adverse events and five are male-related adverse events. The gender disparity in UTIs increased during the pandemic in the elderly but not in adult patients, concordant with evidence that postmenopausal women are most at risk for UTIs [44].

### Gender differences across drug-associated adverse effects

To examine the landscape of gender differences in drug safety, we constructed a network of drug-adverse event associations that are significantly enriched after the onset of the pandemic in women (Figure 4a) and men (Figure 4b). We find that women have 169 enriched associations between drugs and adverse reactions while men only have 51, lending further support to our findings that the pandemic exerted a greater influence on the side effect landscape for women.

**Figure 4:**
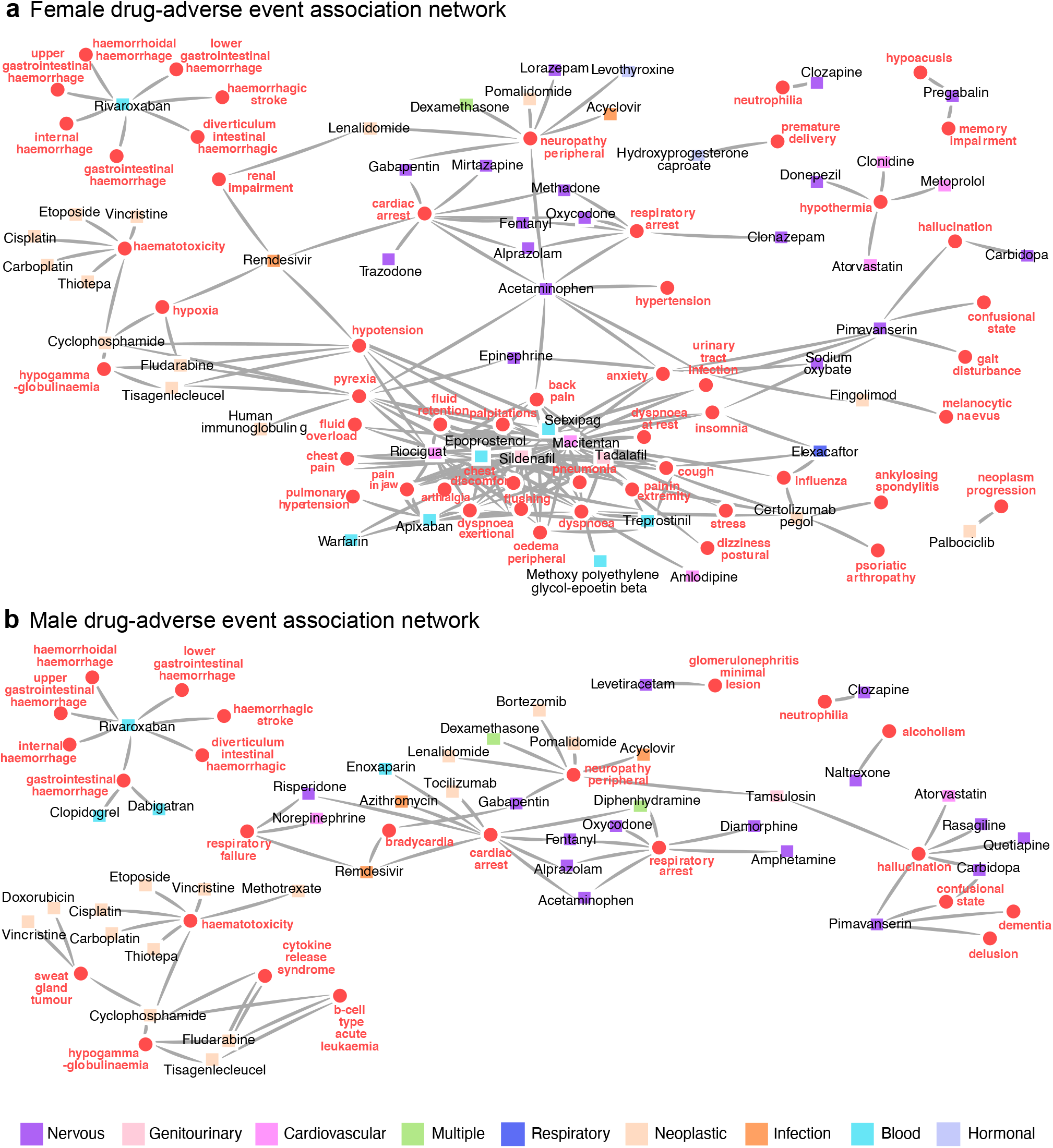
Gender differences in the drug-adverse event association network. Networks constructed from drug-adverse event associations disproportionally enriched in women or men. Nodes represent adverse drug reactions (red circles) and drugs (squares). Node color of drugs is determined by the Anatomical Therapeutic Chemical Classification (ATC) [65], dividing drugs into different groups according to the organ on which they act and their therapeutic, pharmacological and chemical properties. Only nodes with at least one link are shown. Layout is determined by the Fruchterman-Reingold algorithm and manually adjusted for clarity of display. **(a)** Drug-adverse event associations enriched in women during the pandemic more than would have been expected had the pandemic not occurred. **(b)** Drug-adverse event associations enriched in men during the pandemic more than would have been expected had the pandemic not occurred. The network in women has more edges (169 associations) than in men (51 associations), suggesting there are significant levels of drug safety inequities that disproportionately impacted women during the pandemic, compared to men. Associations between cardiac arrest and nervous system drugs are found in both networks. A link between Remdesivir and respiratory failure is found in male but not in female patients.

Several features of the enriched drug-adverse events networks are shared between men and women, such as the cluster of haemotoxicity-associated drugs and the cluster of haemorrhage-related side effects associated with Rivaroxaban. In both networks, cardiac and respiratory arrest are associated with a similar cluster of nervous system-related drugs, including common Benzodiazapines and Opioids such as Fentanyl, Oxycodone, Methadone, and Diamorphine. Many of these drugs are known to be highly addictive and sheds light on how the COVID-19 pandemic has impacted the ongoing substance abuse epidemic [45, 46]. Moreover, Methadone is only enriched in women, while Diamorphine is only enriched in men, suggesting a potential gender difference in the use of addictive substances.

A major difference between the two networks is the large cluster found only in the female network. This cluster includes anxiety and closely associated side effects (such as dizziness, insomnia, palpitations, and dysponea): consistent with numerous studies that women are more likely to report a mental health disturbance during the pandemic [47–49]. Comparing drugs present across the female and male networks is hindered by lacking information on total drug usage in the FAERS dataset. However, both Sildenafil and Tadalafil are located within the center of the female-specific cluster, two drugs whose on-label use is for erectile dysfunction. Despite majority of usage being in men, both drugs are found only the female cluster. Both have off-label uses for treating sexual dysfunction in women, highlighting our method’s ability to detect potential adverse events of off-label drug usage, and to overcome the limitation of differences in drug usage. There are 35 drug-adverse events associations enriched in only male patients. For instance, an association between alcoholism and Naltrexone is enriched in men, but not women, indicating a gender disparity in the previously documented rise in alcohol use during the pandemic [50].

We separately analyze associations with Remdesivir as the first treatment for COVID-19 to receive emergency approval [51]. Remdesivir presents in both male and female networks and shows association with cardiac arrest. Analyzing differential rates of adverse events in patients reporting use of Remdesivir, we note that female patients are at increased risk for hypoxia, hypotension, and renal impairment, while in the male network, Remdesivir is connected to respiratory failure. These adverse events have been reported in Remdesivir clinical trials [52, 53], but are very rare (cardiac arrest, 1/155 patients; renal impairment, 4/53; hypotension, 4/53; respiratory failure 2/53) [54] and the trials fail to provide any differential drug effects for distinct patient subsets; the literature does not reveal the association with hypoxia (ROR=7.05; p-value < 10^−36^, Fisher’s exact test). The detection of Remdesivir-associated adverse events highlights the importance of population-scale patient safety datasets to detect rare adverse events and unexpectedly high-risk demographic subgroups. As more clinical treatments and vaccines receive emergency use authorization, rapidly detecting rare adverse events and stratifying at-risk populations will be critical to patient safety.

## Discussion

Here we demonstrate the effectiveness of population-scale patient safety data in revealing demographic variation of drug adverse events during a public health crisis. Using 10,443,476 reports that span 3,624 approved drugs and 19,193 unique adverse events, our algorithmic approach quantifies reporting patterns within and across diverse patient cohorts to flag at-risk demographics and identifies relationships between adverse events and drugs.

Our analyses quantify associations between adverse events and the pandemic and discovers inequalities across patient subpopulations, paving the way for future research to identify medical and non-medical factors driving the inequities. The approach detects population specific side effects supported by the literature, such as increased susceptibility of women to anxiety related effects and the age and gender disparity in urinary tract infections. It also detects side effects that contradict the literature (e.g., increased susceptibility of the elderly to drug-related psychiatric events, despite documented resilience to psychiatric disorders). The models detect novel, rare adverse events, such as hypoxia as a side effect of Remdesivir, highlighting the role of algorithmic models for pharmacosurveillance of treatments granted emergency approval.

The power to detect subtle patterns of drug safety depends on the algorithm’s ability to exclude potential confounding factors. To correct for well-recognized confounding factors, we directly compare reports submitted in 2020 to those in 2019 and use temporal prediction between 2013 and 2020 to identify drug safety events whose reporting changed during the pandemic more than expected had the pandemic not occurred. Further research is necessary to pinpoint driving factors that made the patient safety landscape so different with the pandemic onset, including altered drug usage [55], limited access to healthcare resources [56], and changes to human behavior (like less physical activity [57]). There is an important limitation to consider in interpreting our findings. The patient safety dataset comprises voluntarily submitted reports that are not necessarily representative of prevalence rates of adverse drug reactions [58]. Further, the pandemic has likely affected reporting rates, which can vary across adverse events and time [9]. Interestingly, the total number of adverse event reports decreased in 2020 relative to 2019. Nevertheless, we find that adverse events whose reporting frequency has changed relative to pre-pandemic levels (Figure 1b) tend to be reported considerably more often than expected based on historical data (Figure 1c). This observation, together with abundant research on clinically relevant insights extracted from patient safety datasets [59–64], further strengthen confidence in our key findings.

Our algorithmic approach can identify differential reporting patterns in patient cohorts formed as a function of gender, age, adverse events, and drugs. With additional information on medical and non-medical patient characteristics such as race and ethnicity, the approach is suitable for systematic safety surveillance to pinpoint individuals at high risk for safety events based on risk-altering interactions. We also present a new resource of adverse drug effects and drug-event associations for use in pharmacoepidemiology and public health policy to inform medication use in diverse populations. We expect this algorithmic approach to enable comparison of the COVID-19 pandemic to other health emergencies (like the nationwide opioid crisis in the U.S. and emergencies resulting from hurricanes and wildfires) to unveil the disruptive nature of public health crises on patient safety.

## Data availability

All data used in the paper, including the raw and processed adverse event report dataset, adverse event ontology, drug ontology, the final and intermediate results of the analyses are shared with research community via the project website at https://zitniklab.hms.harvard.edu/projects/patient-safety. We also deposited the data in the Harvard Dataverse repository, giving our collection of datasets a unique identifier: https://doi.org/10.7910/DVN/G9SHDA.

## Code availability

Python implementation of the methodology developed and used in the study is available via the project website at https://zitniklab.hms.harvard.edu/projects/patient-safety. The code to reproduce results, together with documentation and examples of usage, are at https://github.com/mims-harvard/patient-safety.

## Data Availability

All data used in the paper, including the raw and processed adverse event report dataset, adverse event ontology, drug ontology, the final and intermediate results of the analyses will be shared with research community via https://zitniklab.hms.harvard.eduuponpublication.

https://github.com/mims-harvard/patient-safety

## Acknowledgements

X.Z., M.S., and M.Z. are supported, in part, by NSF grant nos. IIS-2030459 and IIS-2033384, and by the Harvard Data Science Initiative. The content is solely the responsibility of the authors. We thank P. Chandak and H. Nilforoshan for useful comments and discussion.

## Authors contribution

X.Z. retrieved, processed, and analyzed the adverse event reporting dataset. M.S. analyzed drug-AE networks. X.Z., M.S., and M.Z. contributed new analytic tools and wrote the manuscript. M.Z. designed the study.

## Methods

The Methods is structured as follows: 1) description of datasets used, 2) description of the algorithmic approach for constructing population-specific models of patient safety.

## Datasets

### Population-scale patient safety dataset

The adverse events reports used in this work are from the FDA Adverse Event Reporting System (FAERS; https://fis.fda.gov/extensions/FPD-QDE-FAERS/FPD-QDE-FAERS.html): a primary source of post-marketing pharmacovigilance. The reports in FAERS mainly contain demographic information (such as age and sex, no personal identifiers), drugs (drug substances) and adverse events (preferred terms in MedDRA; more details in the section on adverse event identification). We investigate 10,443,476 reports, involving 19,193 adverse events and 3,624 drugs, reported between January 2013 and September 2020. For reports with the same case number, we keep only the latest report. We restrict our analysis to the adverse events occurred in the USA (6,351,817 reports) to alleviate country-wise biases and avoid biases caused by different national surveillance systems. The World Health Organization (WHO) declared a pandemic on March 11, 2020 (retrieved from WHO). The latest available safety reports were submitted on September 30, 2020: thus, we focus on the reports submitted from March 11 to September 30 in 2020 and the same period in previous years (from 2013 to 2019), leading to 3,709,531 reports.

### FDA reporter qualifications

FAERS submissions are voluntarily made by the reporters, who send the reports to the FDA directly or through drug manufacturers. The reporters include health-care professionals (physicians, pharmacists, nurses, dentists, etc.) and non-professionals (lawyers and customers). We investigate the distribution of reporters: physicians (594,787, 16.0%), pharmacists (282,323, 7.6%), other healthcare professionals (548,261, 14.8%), lawyers (113,744, 3.1%), customers (2,045,491, 55.1%), and unknown reporters (124,925, 3.37%). To increase the validity of our analysis, we focus on the reports submitted by healthcare professionals (1,425,371 reports).

### Demographic distribution

We further look into the distribution of diverse patient cohorts and find that the proportion of men, women, and unknown sex are 484,649 (34.0%), 784,230 (55.0%), and 156,492 (11.0%), respectively. Based on the aging criteria set by the WHO [66], we split the patients into young (<20 years, 35,987 reports, 2.52% of all reports, mean=13.9, std=9.0), adults (20∼65 years, 508,983 reports, 35.7%, mean=47.8, std=13.2), elderly (>65 years, 305,685, 21.4%, mean=72.9, std=11.1), and unknown age (574,716 reports, 44.3%). In sex- or age-specific analysis, we omit reports with unknown sex or unknown age, respectively. For instance, we ignore patients unknown sex when calculating the proportion of female patients in adverse event reports (Figure 2b).

### Adverse event identification

The drug reactions recorded in FAERS reports are characterized by preferred terms (PT; text string) in MedDRA ontology ((https://www.meddra.org/how-to-use/basics/hierarchy) [41]. We map the adverse events from their PT string in FAERS to the corresponding MedDRA PT identifier for downstream analysis. One of the major challenges faced by the mapping is that the string names (*e*.*g*., ‘Phobia fear \\ Height’) shown in adverse event reports may not match with the standard MedDRA PT names because of nonstandard annotations. Addressing this issue, we propose a simple but efficient mapping strategy which contains three steps. First, we convert all strings into lower case (*e*.*g*., ‘phobia fear \\ height’); second, split the string by ‘\\’ if exist, and then separately map the split strings (*e*.*g*., ‘phobia fear’ and ‘height’) to MedDRA; third, if none of them match with the standard PT names, we further split the string by ‘‘if exist (*e*.*g*., ‘phobia’ and ‘fear’) and check the matching (*e*.*g*., ‘phobia’ match with MedDRA ID ‘10034912’). In this way, we can map 98.2% of all the adverse events strings appeared in FAERS dataset, which is higher than most literature (such as [67]). We separately analyze reports with explicit references to COVID-19. That included 6 adverse events (COVID-19 (1,674 reports), COVID-19 pneumonia (135 reports), suspected COVID-19 (103 reports), asymptomatic COVID-19 (14 reports), coronavirus infection (547 reports), and coronavirus test positive (103 reports)). When analyzing gender inequality, we treat sex-specific adverse drug reactions separately. For example, when measuring the impact of the pandemic on the gender gap, we manually excluded premature delivery because it only occurs in female patients.

### Mapping of adverse events to human organ systems

To explore the disparities among different parts of human body, we categorize adverse events into higher level, from ‘Preferred Terms (PTs)’ level to ‘System Organ Classes (SOCs)’ level, based on etiology (such as infections and infestations) and manifestation sites (such as gastrointestinal disorders) following the MedDRA hierarchy. We allow selective exclusion before feeding data into the proposed framework. We manually remove the adverse events related to four SOCs (*i*.*e*., social circumstances, surgical and medical procedures, product issues, and investigations) that are not related to medication therapies. Furthermore, to increase the robustness and generalizability of the results, we focus on the adverse events that are observed in at least 100 reports either before or during the pandemic.

### Mapping of drugs, controlled drug vocabulary, and Anatomical Therapeutic Chemical (ATC) classification system

We first map the drugs in the AE reports to DrugBank IDs (https://go.drugbank.com) [68]. We implement the same mapping strategy as described in the section on mapping of adverse events. We also group drugs into categories given by the Anatomical Therapeutic Chemical (ATC) classification system. The ATC categorization is an internationally accepted classification system maintained by the WHO (https://www.whocc.no/atcdddindex) [65] that classifies active ingredients of drugs according to the organ or system on which they act and their therapeutic, pharmacological and chemical properties. For example, drug Ritonavir is annotated with ATC codes J05AR10, J05AP53, J05AE03, J05AR23, J05AR26, and J05AP52, indicating that Ritonavir is an antiviral drug used for treatment of HIV and HCV infections.

### Population-scale adverse event model of patient safety

#### Overview of the approach

The approach receives multimodal information including drugs, adverse events, and demographic (*e*.*g*., gender and age) from adverse event reports in order to identify the substantial drug side effects that are significantly associated with pandemic, and detect the inequality among demographic subpopulations. The approach has three components. (1) We first employ disproportionality estimation to identify the adverse drug reactions that are significantly associated with pandemic. (2) We then track the evolving trace of each adverse event between 2013 and 2019 and quantify its expected incidence proportion in 2020. We remove the adverse events whose abnormal reporting frequency in the pandemic can be explained by their temporal trend. (3) Finally, we remove drug confounders by considering two kinds of drug interference: we only keep the adverse events that are significantly associated with at least one drug and the formed drug-adverse event pairs are significantly associated with the pandemic.

Next, we take the overall population (SI Table S1) as an example to introduce the pipeline of proposed framework. Our model can be flexibly generalized to any demographic subpopulations (such as SI Table S2-S6) by adjusting the input reports. We start with the notation and mathematical representation of the dataset, and then describe details of the three components.

#### Notation and representation of adverse event data

We denote the FAERS dataset as *X* where each element ***x***_*i*_ represents a single patient safety report. We use *D* and *S* to denote the set of all drugs and adverse events appeared in FAERS dataset, respectively. We regard each report as a tuple including a set of drugs *𝒟*_*i*_, a set of adverse events *S*_*i*_, patient’s age *a*_*i*_, sex *g*_*i*_, weight *w*_*i*_, reporter’s qualification *q*_*i*_, severity vector ***b***_*i*_, and reporting date *t*_*i*_; in other words, we have ***x***_*i*_ = (*𝒟*_*i*_, *S*_*i*_, *a*_*i*_, *g*_*i*_, *w*_*i*_, *q*_*i*_, ***b***_*i*_, *t*_*i*_). The patient may take several medications at the same time and have multiple adverse drug events. Thus, each report contains a drug set *𝒟*_*i*_, which is a subset of *𝒟*, and each drug *d*_*j*_ *∈ 𝒟*_*i*_ is represented by its DrugBank ID (string). Similarly, the adverse events set *S*_*i*_ *⊆ S* contains one or more drug side effects and each *s*_*h*_ *∈ S*_*i*_ is represented by its MedDRA ID (string). The patient’s age (in year) *a*_*i*_ is represented by an integer; the biological sex is denoted by *g*_*i*_ *∈ {*1, 2*}* where 1 denotes male and 2 denotes female; the weight (in kilogram) *w*_*i*_ is represented by real number. The reporter’s healthcare qualification *q*_*i*_ *∈ {*1, 2, 3, 4, 5*}* falls in one of the five categories of physicians, pharmacists, other professionals, lawyers and customers (denoted by one to five). The severity vector ***b***_*i*_ is a binary vector with six elements, where 1 denotes severity and 0 denotes not, corresponding to six outcomes (death, life-threatening condition, hospitalization, disability, congenital anomaly, and other medical conditions) of patient. We represent the reporting date *t*_*i*_ by the number of days between the date when the report is submitted and a defined calibration date (January 1, 2000). All the introduced components are used for analysis. For instance, we leverage all the available information (such as demographic data, severity, and submitting date) for patient matching [69–71] in drug interference analysis.

We denote the set of reports submitted in year *k* as *𝒳*_*k*_ where *k ∈ {*2013, …, 2020*}*. The union of every year’s reports equals to the whole FAERS dataset: 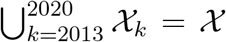. In contrast to most traditional post-marketing studies which only pay attention to specific or few medica tions/reactions [3, 72], our research is conducted in a more complex context, involving the time dimension of patient safety data and investigating a large number of drugs/adverse events.

To better organize the complex multimodal information, we define logical conditions, which allows us to form a cohort of reports as a function of drugs, adverse events and submitting time. A logical predicate *L* consists of a sequence of atomic formulas: drug *d*_*j*_, adverse event *s*_*h*_, and year *k*, which are connected with logical connectives: negation (“not” or *¬*), logical conjunction (“and” or *∧*), logical disjunction (“or” or *∨*), existential quantification (*∃*), and universal quantification (*∀*). We use “*·*” to denote free/unbound variables. As an example, a logical predicate *L* = (*¬d*_*j*_, *s*_*h*_, *·*) denotes the following conjunctive connection: “Report describes a patient who does not take drug *d*_*j*_ <u>and</u> Report indicates occurrence of adverse event *s*_*h*_ <u>and</u> Report is submitted anytime in 2013-2020 time window.” We define *f* (*L*) as a function of *L* that selects all adverse event reports that satisfy logical predicate *L*. We formulate the value of *f* as:

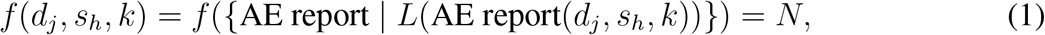

where *N* represents the number of AE reports that satisfy the atomic formulas of drug *d*_*j*_, adverse event *s*_*h*_, and submission time *k* connected by logical conjunction. Let’s look at an example: *f* (Pimavanserin, hallucination, 2020) = 1,145 selects a set of reports for which the following holds: “A patient received Pimavanserin treatment <u>and</u> later experienced unwanted side effect of hallucination, <u>and</u> this adverse drug reaction was submitted to the FDA in 2020 (Mar 11 – September 30).” In this particular case, there are 1,145 patients in the FAERS who meet the above requirements.

Our approach receives the adverse event data *𝒳* and identifies adverse events *𝒮*^*′*^ (*𝒮* ^*′*^ *⊆ 𝒮*), where each drug side effect 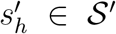 has a significantly different reporting pattern during the pandemic than would have been expected had the pandemic not occurred. To that end, we define three reporting odds ratios (RORs [73–75]): *β*(*s*_*h*_) measures the association between adverse event *s*_*h*_ and the pandemic (SI Table S7); *γ*(*d*_*j*_, *s*_*h*_) quantifies the connection between drug *d*_*j*_ and adverse event *s*_*h*_ (SI Table S8); *δ*(*d*_*j*_, *s*_*h*_) estimates the association between drug-adverse event pair (*d*_*j*_, *s*_*h*_) and the pandemic (SI Table S9).

#### Step 1) Disproportionality estimation

We conduct the disproportionality estimation [67, 76] on each adverse event to examine the association between the adverse event and pandemic (March 11–September 30, 2020) in contrast to before the pandemic (March 11–September 30, 2019). While disproportionality analysis is an established approach for pharmacovigilance to generate hypotheses on possible causal relations between drugs and adverse effects [72, 77], we here use it in a novel way that quantifying the association between adverse events and their submission periods. For each *s*_*h*_ *∈ 𝒮*, we define *β*(*s*_*h*_) to measure the strength of association between *s*_*h*_ and pandemic by comparing the reporting frequency during the pandemic with it before the pandemic. Taking *s*_*h*_ as input, we calculate *β*(*s*_*h*_) as:

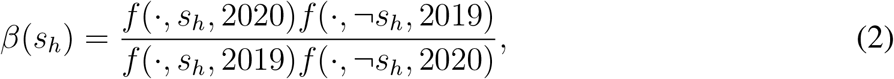

where *f* (*·, s*_*h*_, 2020) and *f* (*·, s*_*h*_, 2019) represent the number of reports involving *s*_*h*_ in March 11 to September 30 in 2020 and 2019, respectively; *f* (*·, ¬s*_*h*_, 2020) and *f* (*·, ¬s*_*h*_, 2019) denote the number of reports which not contain *s*_*h*_ in March 11 to September 30 in 2020 and 2019, respectively.

We quantify the upper/lower 95% confidence interval (CI) of *β*(*s*_*h*_) 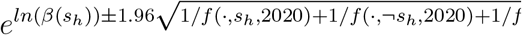 Table S7). This calculation does not consider the constraint of medication.

We calculate the significance values using the Fisher’s exact test followed by the Bonferroni correction for multiple hypothesis testing. We keep only the adverse events, which pass both the significance test (adjusted p-value < 0.05) and the ROR criterion. Regarding *β*(*s*_*h*_), the ROR-based selection criterion is that the lower 95% CI is greater than 1 for adverse events that are reported more frequently during the pandemic (enriched or overrepresented); or that the upper 95% CI is less than 1 for adverse events that are reported less often during the pandemic (purified or underrepresented). Unlike previous studies that mainly focused on drug responses with higher ROR (such as ROR > 1) [31], our model able to detect both overrepresnted (*β*(*s*_*h*_) > 1) and underrepresented (*β*(*s*_*h*_) < 1) adverse events.

#### Step 2) AE reporting trajectories

The reporting trajectory of an adverse event refers to the changing trend of adverse event incidence proportion, indicated by its temporal/historical data. For example, if the reporting frequency of a certain adverse drug reaction has continually increased from 2013 to 2019, it is not surprise that it also grows from 2019 to 2020, and we cannot assertively attribute its overrepresentation to the pandemic. Addressing this issue, we propose a dedicated indicator, pandemic-adverse event association index (PAEAI), to measure whether a medication’s incidence conforms to its trajectory. We next report how to reduce the confounding factor of temporal trend in single adverse event level.

We regard March 11–September 30 in 2020 as pandemic period and in previous years (2013 to 2019) as non-pandemic periods. For *s*_*h*_ which pass the two-fold criterion in previous step, we build its trajectory vector ***v***(*s*_*h*_) = [*v*_*h*,2013_, *v*_*h*,2014_, …, *v*_*h*,2020_]. Element *v*_*h,k*_ represents the pro-portion of reports related to *s*_*h*_ in all reports submitted in year *k*. For example, of the 211,152 reports submitted during the pandemic period, 6,130 involve hallucination, which means the proportion of hallucination during the pandemic is 2.9% = 6, 130*/*211, 152. Inspired by the powerful temporal feature capture ability of autoregressive method [78], we train a second-order autoregressive regression (*i*.*e*., AR(2)) model for each adverse event by fitting its historical values [*v*_*h*,2013_, *v*_*h*,2014_, …, *v*_*h*,2019_]. The regression models trained on different adverse events do not share parameters. The optimized model is then used to predict the proportion of *s*_*h*_ in each year. The predictions 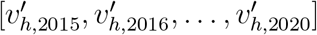 is from 2015 to 2020 since 2-order AR model need the first two numbers (in 2013 and 2014) as initial inputs. On top of the difference between observation and prediction, we define PAEAI of *s*_*h*_ as:

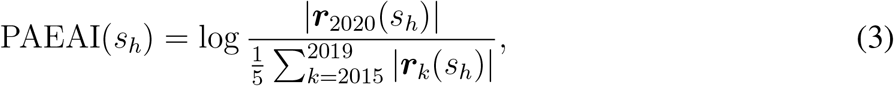

where ***r***_*k*_(*s*_*h*_) denotes the standardized residual in year *k* in the regression model of *s*_*h*_, which is calculated through 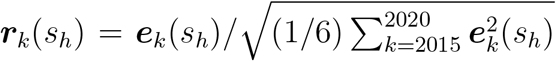. The 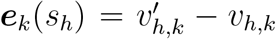 denotes the residual of *s*_*h*_ in year *k* (from 2015 to 2020).

In summary, PAEAI measures the ratio of average standardized residual in the pandemic relative to pre-pandemic. Positive PAEAI indicates the reporting frequency of *s*_*h*_ has changed during the pandemic more than would have been expected had the pandemic not occurred. It suggest the adverse event *s*_*h*_ is associated with the pandemic factors and cannot be explained by temporal trends based on historic data. Negative PAEAI indicates the change during the pandemic does not exceed expected normal fluctuations of the *s*_*h*_’s trajectory. A higher value of PAEAI indicates more substantial changes in the reporting frequency of *s*_*h*_. As PAEAI takes the logarithm, a small difference in PAEAI reflects a rather large change during the pandemic relative to prepandemic levels. In the next step of the approach we only consider *s*_*h*_ with positive PAEAI.

#### Step 3) Drug interference

Next, we reduce the confounding effects of multiple drugs. Traditional pharmacosurveillance typically only concern how adverse events connected to specific therapy [32], however, our approach able to discover multiple drugs that may explain the change of reporting frequency during the pandemic, and control underlying variables in analysis. We consider two types of medication interference:

- The first interference is that the adverse event co-occur with a certain drug in safety reports but their association may not significant.
- The second interference is that the association between an adverse event and the pandemic can be attributed to multiple drugs, however, none of the formed drug-adverse event pairs is significantly associated with the pandemic.

Accordingly, we consider two aspects to prevent drug interference. First, the adverse event (such as hallucination) should be significantly associated with the therapy of at least one drug (like Pimavanserin). Second, the formed drug-adverse event pair (like Pimavanserin-hallucination) should be significantly associated with the pandemic.

To eliminate the first interference, for a certain adverse event *s*_*h*_ that has passed the selection in previous two components, we go through all the drugs that co-occurred with it in adverse event reports during the pandemic (*i*.*e*., *𝒳* _2020_). We use 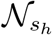 to denote the set of found drugs for *s*_*h*_. We only check association between *s*_*h*_ and 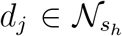 during the pandemic, for simplification, we omit the subscript of year while analyzing the first medication interference. For a specific drug 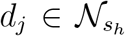, we regard the reports where the drug is involved as positive samples 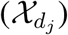, while not involved as negative samples 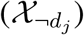. All the positive samples are included in a test group 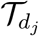. To keep the comparison fair, we select a subset from negative samples as control group 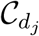 where the patients have similarly situation with the ones from 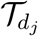(*i*.*e* 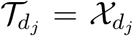 and 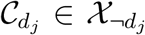) [67, 79]. The reports in 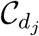 are selected through a nearest neighbour propensity score matching model [79] which measures the similarity between a report (a patient) from negative samples with a report from positive samples as a function of the patient characteristics. Based on the available information, we build a characteristic factor ***z***_*i*_ = [*a*_*i*_, *g*_*i*_, *w*_*i*_, *q*_*i*_, *b*_*i*,1_, …, *b*_*i*,6_, *t*_*i*_] for each report, including the patient’s age, sex, weight, the qualification of the reporter, severity vector (*b*_*i*,1_ to *b*_*i*,6_ are the six elements in ***b***_*i*_) and the submission date. For each report in test group 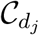, we select 10 reports that have the highest propensity scores into control group [67], which makes 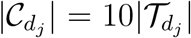. The propensity scores are measured by the cosine similarity among characteristic factor. Afterwards, based on 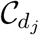 and 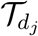, we define *γ*(*d*_*j*_, *s*_*h*_) for each *s*_*h*_ and all it’s co-occurred drug 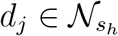 by:

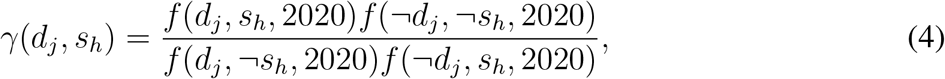

where *f* (*d*_*j*_, *s*_*h*_, 2020) represents the quantity of reports that exposed to a certain drug *d*_*j*_ and mean-while emerged the specific drug reaction *s*_*h*_, whereas *f* (*d*_*j*_, *¬s*_*h*_, 2020) denotes the number of reports that also exposed to *d*_*j*_ but not emerged *s*_*h*_; *f* (*¬d*_*j*_, *s*_*h*_, 2020) represents the number of reports that not involves *d*_*j*_ but generated *s*_*h*_, while *f* (*¬d*_*j*_, *¬s*_*h*_, 2020) denotes the amount of reports that not involves *d*_*j*_ and not contain *s*_*h*_. We quantify the upper/lower 95% CI of *γ*(*d*_*j*_, *s*_*h*_) by 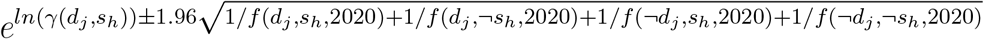 (SI Table S8).

The passing requirements are the 95% CI of *γ*(*d*_*j*_, *s*_*h*_) not cross one and the p-value corrected by Bonferroni method smaller than 0.05. To this end, for each *s*_*h*_, we have a drug set 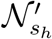 which is a subset of 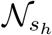, where each drug 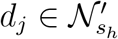 is significantly associated with *s*_*h*_.

To alleviate the impact of the second interference, we further explore whether each drug-adverse event pair is significantly related to the pandemic in contrast to before the pandemic. We only consider *s*_*h*_ that is significantly associated with at least one drug (*i*.*e*., 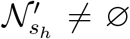). We then define *δ*(*d*_*j*_, *s*_*h*_), denoting the odds ratio of pair (*d*_*j*_, *s*_*h*_) (where 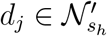) during the pandemic,

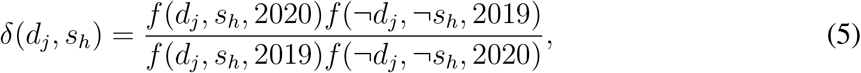

where *f* (*d*_*j*_, *s*_*h*_, 2020) and *f* (*d*_*j*_, *s*_*h*_, 2019) denote the number of reports that contains the drug-adverse event pair of interest in 2020 and 2019, respectively; *f* (*¬d*_*j*_, *¬s*_*h*_, 2020) and *f* (*¬d*_*j*_, *¬s*_*h*_, 2019) denote the volume of reports that not contains the (*d*_*j*_, *s*_*h*_) pair in 2020 and 2019, respectively. We calculate the upper/lower 95% CI of *δ*(*d*_*J*_, *s*_*h*_ through 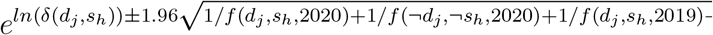 Table S9). The criterion of significance is the same as the previous stage (i.e., 95% CI of *δ*(*d*_*j*_, *s*_*h*_) not cross one and Bonferroni adjusted p-value < 0.05). Thus, for *s*_*h*_, we have a set of drugs, de-noted by 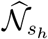 and 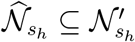, where each drug 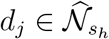 satisfies that the drug-adverse event pair (*d*_*j*_, *s*_*h*_) is significantly associated with the pandemic. At last, any adverse event 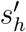 with 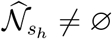 satisfies that its changed reporting frequency can be attributed to at least one medication, and the drug-adverse event pair is significantly associated with the pandemic.

In summary, each adverse event 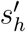 identified by our approach have multiple defined metrics describing different aspects. The *β* (along with 95% CI and adjusted p-value) detects the reporting frequency during the pandemic, which is above (overrepresented) or under (underrepresented) what we expected (see the section on disproportionality estimation). The PAEAI guarantees the abnormal reporting frequency in 2020 can not be explained by its temporal trend from 2013 to 2019 (see section on AE reporting trajectories). Each 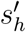 has one or more *γ* and *δ* (and corresponding 95% CIs and adjusted p-values) to ensure that the changed reporting frequency during the pandemic is not affected by drug interference (see the section on drug interference).

