## Supplementary Table 1-9, Figure 1-9 for "Population-scale patient safety data reveal inequalities in adverse events before and during COVID-19 pandemic"


This PDF file includes:

Supplementary Figures 1 to 9

Supplementary Tables 1 to 9

#### Contents

#### List of Figures


S1 Number of identified adverse events across demographic groups. . . . . . . . . . . . . S3
S2 Distribution of enriched adverse events by organ type and demographic group. . . . . S4
S3 Distribution of purified adverse events by organ type and demographic group. . . . . S5
S4 Gender gap in all patients normalized by population size. . . . . . . . . . . . . . . . . S6
S5 Gender gap in adults normalized by population size. . . . . . . . . . . . . . . . . . . . S7
S6 Gender gap in elderly normalized by population size. . . . . . . . . . . . . . . . . . . S8
S7 Comparison of proportion of male patients in enriched adverse events before and during the pandemic. . . . . . . . . . . . . . . . . . . . . . . . . . . . . . . . . . . S9
S8 Comparison of reporter distribution in purified adverse events before and during the pandemic (continued in Figure S9). . . . . . . . . . . . . . . . . . . . . . . . . . S10
S9 Comparison of reporter distribution of purified adverse events before and during the pandemic (continued). . . . . . . . . . . . . . . . . . . . . . . . . . . . . . . . . . S11


#### List of Tables


S1 Summary of adverse events identified by our approach in all patients. . . . . . . . . . S12
S2 Summary of adverse events identified by our approach in male patients. . . . . . . . S14
S3 Summary of adverse events identified by our approach in female patients. . . . . . . S15
S4 Summary of adverse events identified by our approach in young patients. . . . . . . . S17
S5 Summary of adverse events identified by our approach in adult patients. . . . . . . . S17
S6 Summary of adverse events identified by our approach in elderly patients. . . . . . . S19
S7 Associations between adverse events and the pandemic. . . . . . . . . . . . . . . . . . S20
S8 Associations between adverse events and drugs. . . . . . . . . . . . . . . . . . . . . . S23
S9 Associations between drug-adverse event pairs and the pandemic. . . . . . . . . . . . S26

### Supplementary Figures

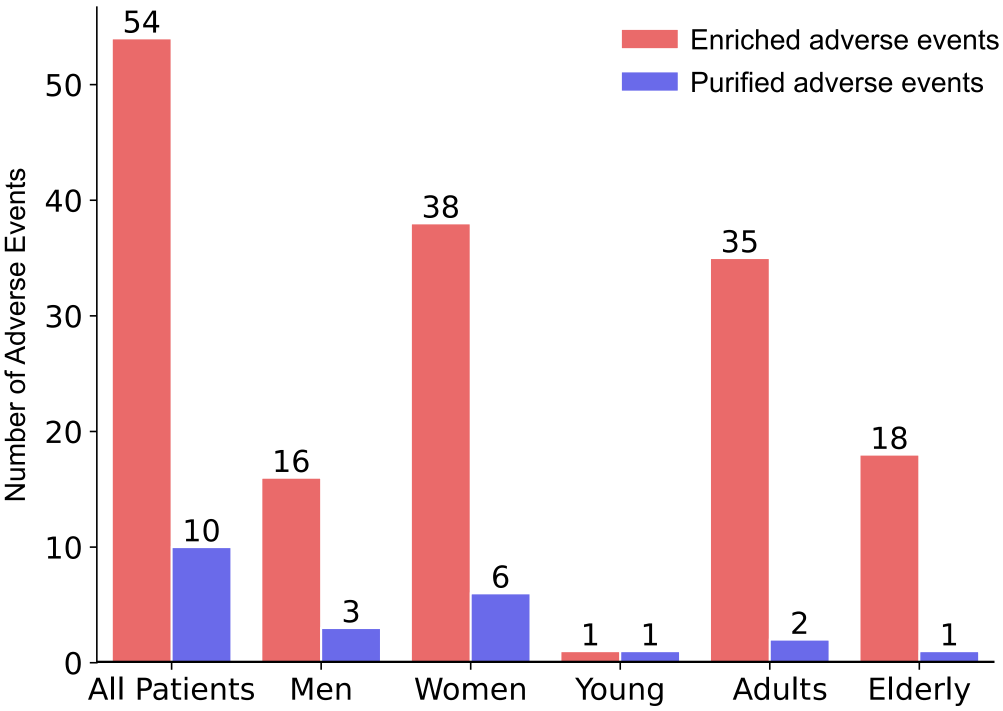


**Figure S1: Number of identified adverse events across demographic groups.** Enriched adverse events refer to the drug side effects that are reported more often during the pandemic compared to before the pandemic, while purified ones are less reported. Although the total amount of reports submitted in the pandemic period (March 11 – September 30, 2020) has dropped to 211,152 in contrast to 220,920 before the pandemic (March 11 – September 30, 2019), we find that the enriched adverse drug reactions are more than, if not equal to, the purified ones. In terms of all patients, the number of enriched adverse events is 5.4 times more than the purified adverse events. This disparity is consistently observed across demographic groups including men (5.33×), women (6.33×), adults (17.5×), and elderly (18×). We find female patients suffer from more enriched adverse events than male patients; adults undergo more than young and aged individuals. Adjusted by the size of populations (per million patients), we find 48.5 adverse events with higher reporting frequency in response to the pandemic in women, whereas only 33.0 in men; there are only 27.8 enriched adverse drug reactions in young people, which is smaller than its counterparts in grow-ups (68.8) and older patients (58.9).

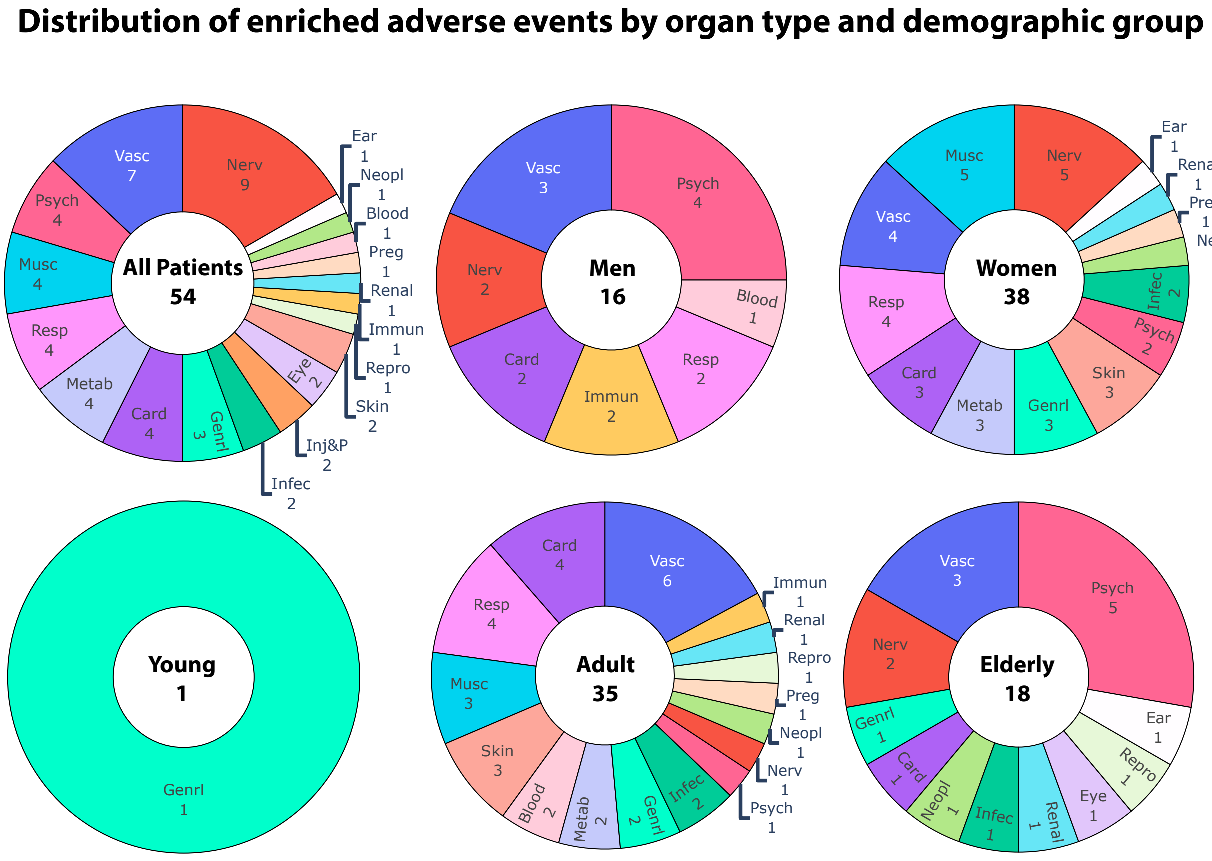


**Figure S2: Distribution of enriched adverse events by organ type and demographic groups.** The colors represent organ types (based on system organ class in MedDRA hierarchy; Methods) and the legend is the same as in Figure 2a. We find the enriched adverse events distribute unequally across organ systems. For example, psychology-related side effects are commonly found in male patients but not in female patients. Our model also detects the different distribution between the overall population and diverse patient cohorts. There are nine adverse events related to the nervous system regarding all patients, however, only one in adults and two in the elderly.

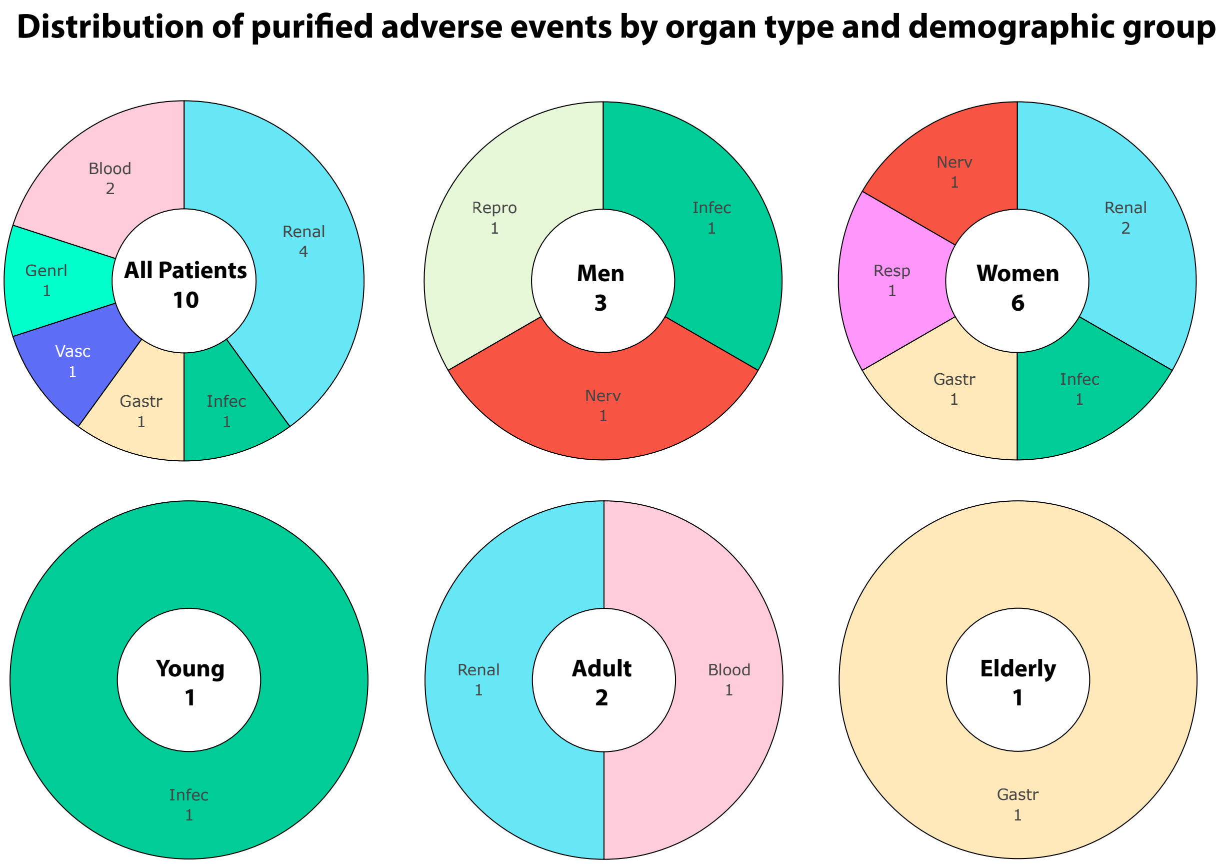


**Figure S3: Distribution of purified adverse events by organ type and demographic groups.** The legend is the same as in Figure 2a where color represents organ systems according to the system organ class (SOC) in MedDRA ontology. This figure presents the adverse events, identified by our framework, which is triggered less frequently during the pandemic, only consider the reports submitted by healthcare professionals (Methods). For example, we find 4 out of 10 purified adverse events in all patients are related to renal disorders, one possible reason is that the diagnosis of renal adverse events highly depends on professional medical facilities which are insufficient during the pandemic. To further take into account the cases reported by nonprofessionals, we investigate the distribution of reporters' qualification of each purified adverse events (Figure S8; Figure S9).

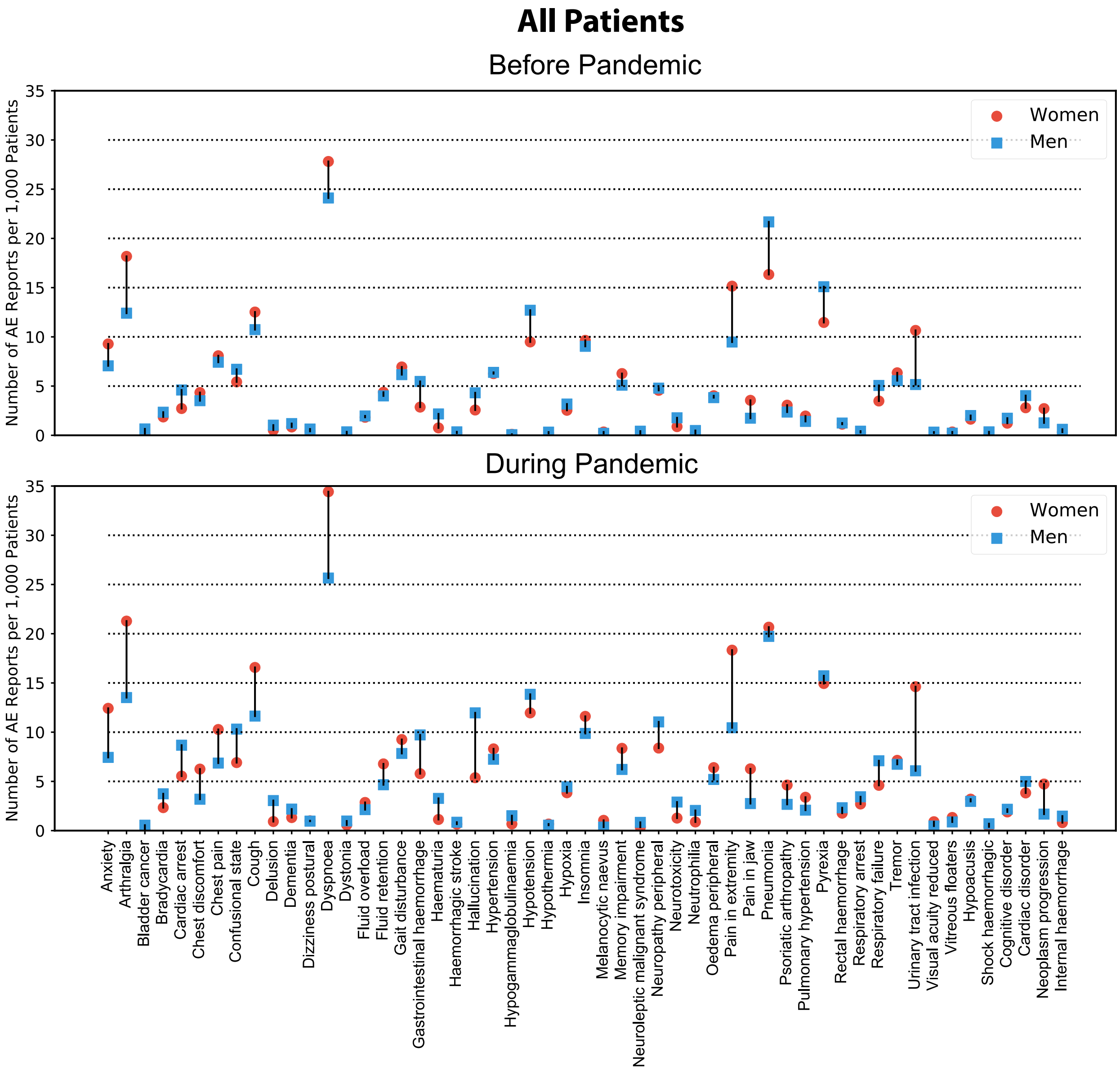


**Figure S4: Gender gap in all patients normalized by population size.** We provide the incidence proportion (*i.e.*, the number of reports regarding a specific adverse event in 1,000 patients) of the enriched adverse events. This figure corresponds to Figure 2c (in the main text) whose y-axis shows the absolute number of reports. We find that gender disparity is increased in most adverse events (62.3%) during the pandemic compared to before the pandemic.

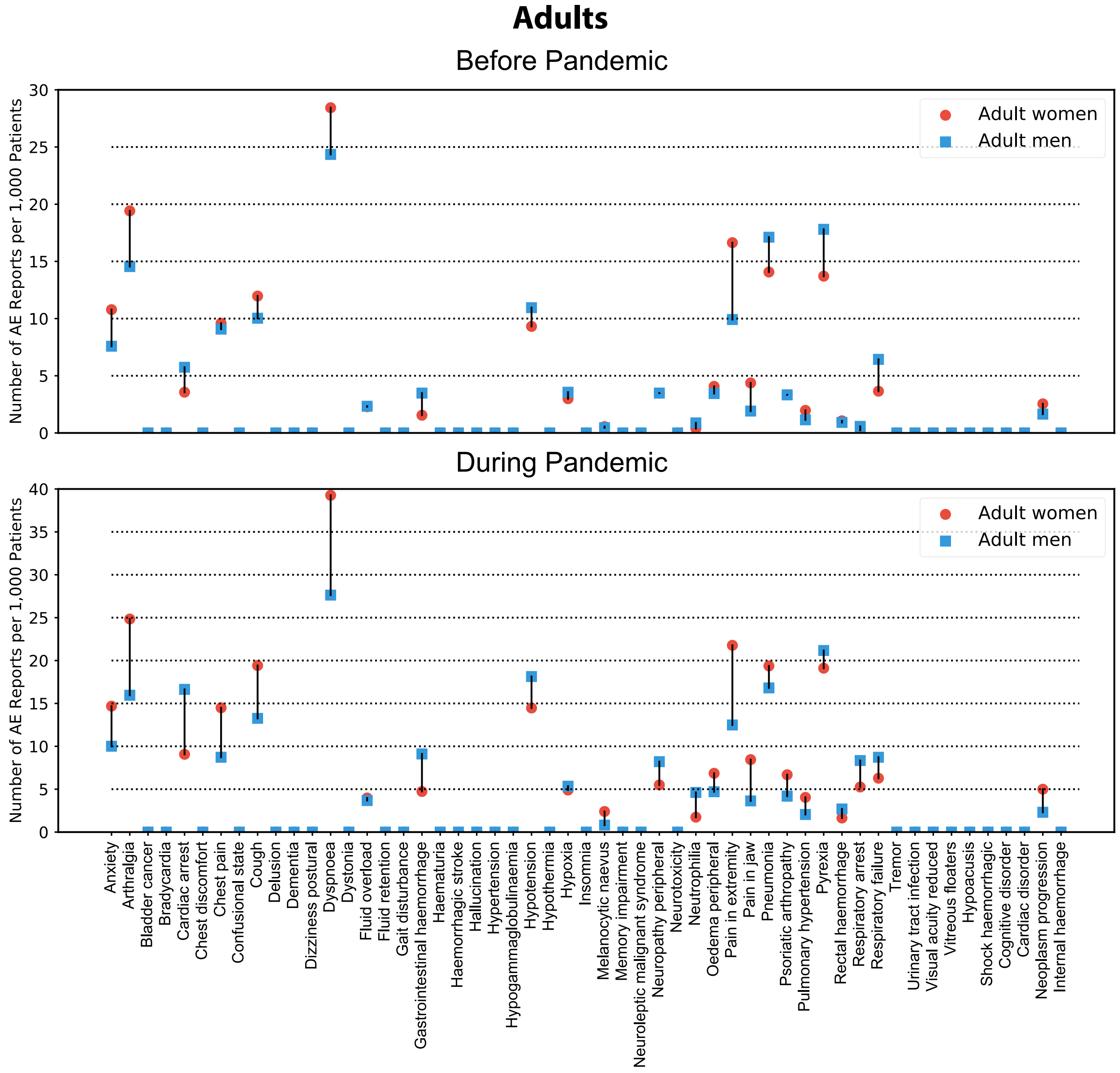


**Figure S5: Gender gap in adults normalized by population size.** Corresponding to Figure 3c that shows the absolute number of reports, the y-axis in this figure presents the relative number of reports adjusted by the number of adult patients (before and during the pandemic, respectively). For better comparison with the counterparts in all patients, we provide all adverse events as shown in the Figure S4. The adverse events with zero reports per thousand adults are not significantly associated with the pandemic in adults (although significant in the overall population), or are only observed in unknown sex. Among 24 drug reactions with gender disparities, we find that 17 experience a larger gender gap during the pandemic.

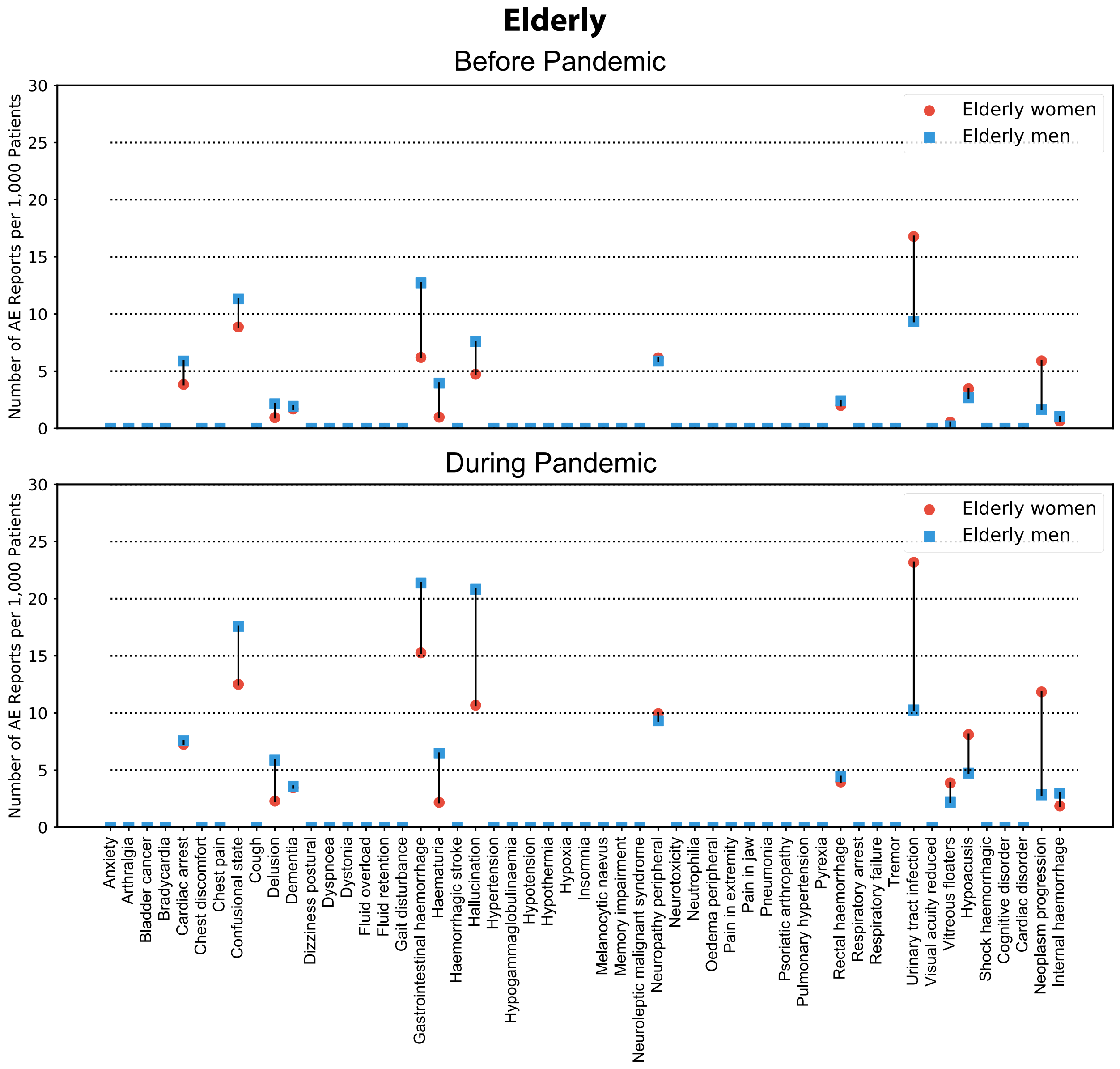


**Figure S6: Gender gap in elderly normalized by population size.** This figure corresponds to Figure 3d with different y-axis (the y-axis in this figure presents the relative number of reports adjusted by the number of elderly patients (before and during the pandemic, respectively)). For better comparison with the counterparts in all patients, we provide all adverse events as shown in Figure S4. The adverse events with zero reports per thousand patients are not significantly associated with the pandemic in elderly (although significant in terms of all patients), or are only observed in unknown sex. We find most (78.6%) of pre-exist gender gaps are increased during the pandemic. For example, before the pandemic, there are 11.33 reports involving confusional state in each thousand female patients, which is 2.47 more than its counterpart in male patients (8.86); the gender difference grows to 5.08 during the pandemic (17.58 in men; 12.5 in women).

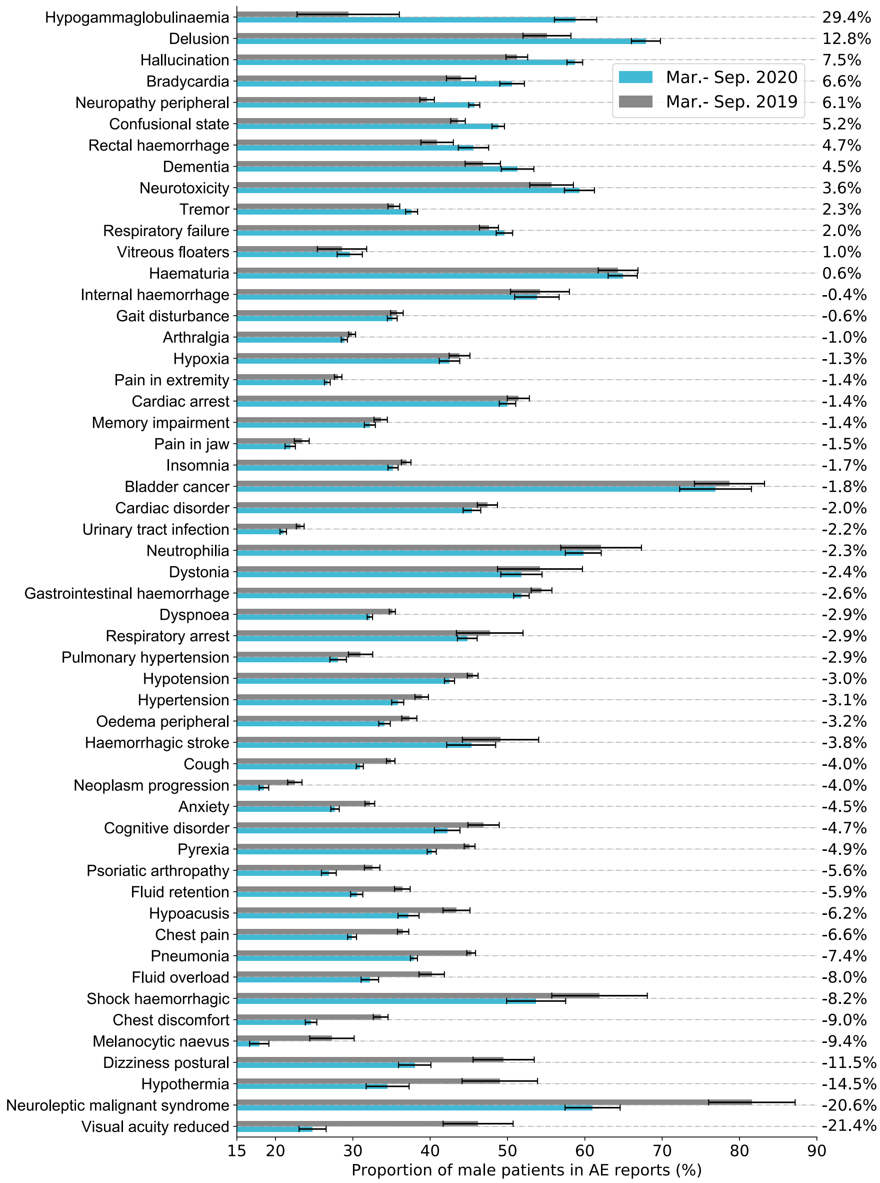


**Figure S7: Comparison of proportion of male patients in enriched adverse events before and during the pandemic.** We present the percentage of men (omit unknown sex) in adverse event reports, corresponding to Figure 2b which shows the proportion of female patients. The annotation shows the increased magnitude during the pandemic (March 11 – September 30, 2020) compared to before the pandemic (March 11 – September 30, 2019). We find the proportion of male patients are increased in 13 out of 53 enriched adverse events during pandemic period. For example, among the reports contains hypogammaglobulinaemia as drug side effect, there are only 29.5% occurred in men before the pandemic but jumped to 58.9% after pandemic onset, claiming an increased margin of 29.4%.

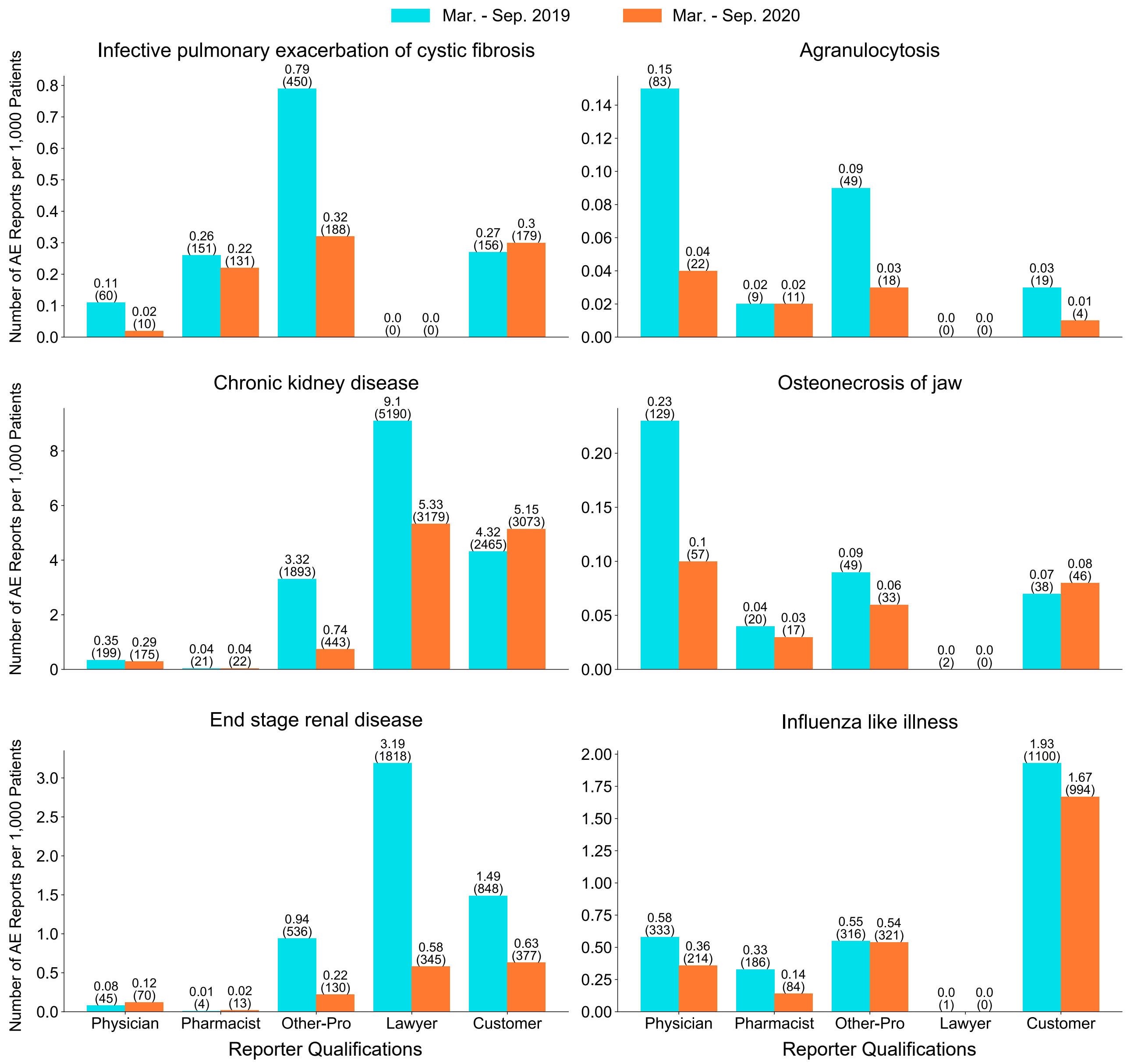


**Figure S8: Comparison of reporter distribution in purified adverse events before and during the pandemic (continued in Figure S9).** The adverse event reports in FAERS are submitted by five categories of reporters: physician, pharmacist, other professionals (such as nurses), lawyer, and customer. Based on the reports submitted by healthcare professionals (the first three qualifications), our approach identifies 10 purified adverse events in all populations. Here, we provide fine-grained reporter distribution by extending to nonprofessional reporters (the last two qualifications). The y-axis shows the number of reports per 1,000 patients and the annotations in brackets present the absolute number of reports.

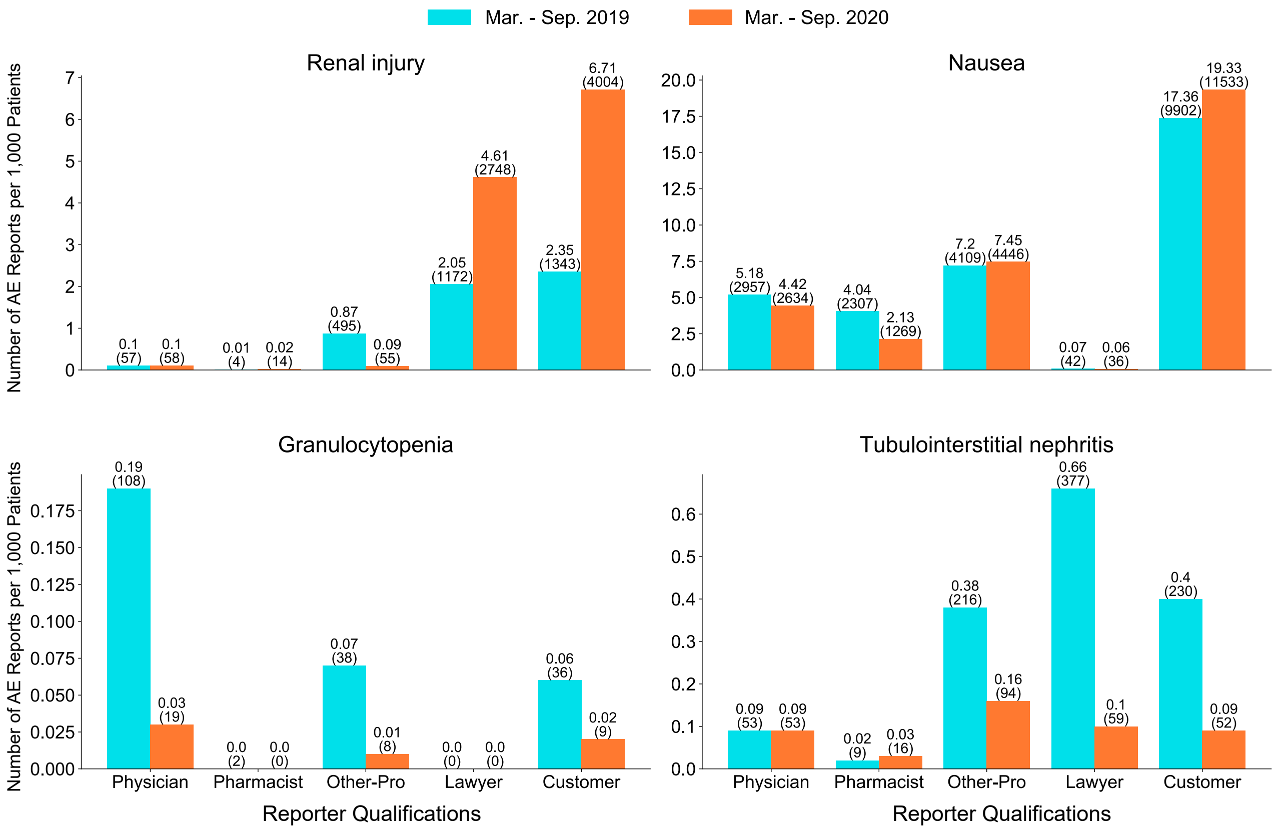


**Figure S9: Comparison of reporter distribution of purified adverse events before and during the pandemic (continued).** Although the incidence proportion are decreased in reports submitted by professionals, we find five adverse events (infective pulmonary exacerbation of cystic fibrosis, chronic kidney disease, osteonecrosis of jaw, renal injury, and nausea) have more self-reported cases (submitted by customer themselves) during the pandemic in contrast to before pandemic. In particular, reports of kidney injury have dropped dramatically among professionals, but have risen sharply among non-professionals, which indicates that patients suffer from kidney injury do not have sufficient professional medical resources during the pandemic.

### Supplementary Tables

**Table S1: Summary of adverse events identified by our approach in all patients.** Shown are 64 adverse events (54 enriched and 10 purified) are identified in the overall population by the proposed framework (Figure 1.a). This table shows the name of adverse events and their corresponding identifier in MedDRA ontology (https://www.meddra.org/how-to-use/basics/hierarchy). The reporting odds ratio $\beta$ represents the association between adverse event and the pandemic (Fisher's exact test; p-value is adjusted by Bonferroni correction; Methods). The numbers in square brackets show the range of lower to upper 95% confidence interval. Higher $\beta$ and smaller p-value indicate the adverse event has stronger association with the pandemic. The defined PAEAI presents the extent (higher PAEAI suggests larger deviation) that adverse event deviates from its historical trend (Methods). All identified adverse events have positive PAEAI which indicates the reporting pattern during the pandemic doesn't follow it's trajectory. The adverse events are sorted in descending order of PAEAI. The 'E' and 'P' in the 'E\P' column denotes this adverse event is enriched or purified, respectively. PEx: pulmonary exacerbations.

| Adverse event | MedDRA ID | $\beta$ (95% CI) | p-value | PAEAI (↓) | E\P |
|---|---|---|---|---|---|
| Delusion | 10012239 | 2.53 [2.08,3.07] | $< 1.6 \times 10^{-18}$ | 1.95 | E |
| Hypothermia | 10021113 | 2.44 [1.79,3.33] | $< 4.0 \times 10^{-5}$ | 1.67 | E |
| Internal haemorrhage | 10075192 | 2.47 [1.92,3.18] | $< 2.0 \times 10^{-9}$ | 1.64 | E |
| Respiratory failure | 10038695 | 1.36 [1.24,1.49] | $< 1.0 \times 10^{-7}$ | 1.58 | E |
| Hypogammaglobulinaemia | 10020983 | 7.68 [5.14,11.48] | $< 4.0 \times 10^{-31}$ | 1.54 | E |
| Bradycardia | 10006093 | 1.38 [1.22,1.56] | $< 1.8 \times 10^{-3}$ | 1.42 | E |
| Shock haemorrhagic | 10049771 | 2.50 [1.76,3.55] | $< 1.0 \times 10^{-3}$ | 1.40 | E |
| Neutrophilia | 10029379 | 2.54 [2.02,3.20] | $< 7.6 \times 10^{-13}$ | 1.32 | E |
| Neuroleptic malignant syndrome | 10029282 | 2.52 [1.77,3.58] | $< 7.3 \times 10^{-4}$ | 1.30 | E |
| Infective PEx of cystic fibrosis | 10070608 | 0.52 [0.46,0.59] | $< 2.6 \times 10^{-19}$ | 1.28 | P |
| Influenza like illness | 10022004 | 0.77 [0.70,0.86] | $< 1.4 \times 10^{-2}$ | 1.26 | P |
| Granulocytopenia | 10018687 | 0.19 [0.13,0.29] | $< 2.4 \times 10^{-16}$ | 1.25 | P |
| Haemorrhagic stroke | 10019016 | 2.44 [1.80,3.29] | $< 1.4 \times 10^{-5}$ | 1.24 | E |
| Dementia | 10012267 | 1.68 [1.41,2.00] | $< 3.5 \times 10^{-5}$ | 1.24 | E |
| Melanocytic naevus | 10027145 | 2.78 [2.04,3.79] | $< 8.7 \times 10^{-8}$ | 1.23 | E |
| Dystonia | 10013983 | 2.51 [1.88,3.34] | $< 7.0 \times 10^{-7}$ | 1.23 | E |
| Dizziness postural | 10013578 | 2.05 [1.61,2.62] | $< 3.6 \times 10^{-5}$ | 1.22 | E |
| Premature delivery | 10036595 | 1.82 [1.49,2.21] | $< 2.1 \times 10^{-5}$ | 1.20 | E |
| Hallucination | 10019063 | 2.42 [2.21,2.65] | $< 5.7 \times 10^{-88}$ | 1.18 | E |
| Hypoxia | 10021143 | 1.48 [1.33,1.64] | $< 8.0 \times 10^{-9}$ | 1.13 | E |
| Chronic kidney disease | 10064848 | 0.31 [0.29,0.34] | $< 2.4 \times 10^{-165}$ | 1.12 | P |
| Pain in jaw | 10033433 | 1.74 [1.57,1.93] | $< 4.9 \times 10^{-22}$ | 1.10 | E |
| Vitreous floaters | 10047654 | 3.85 [2.91,5.10] | $< 3.1 \times 10^{-21}$ | 1.09 | E |
| Pulmonary hypertension | 10037400 | 1.51 [1.32,1.72] | $< 5.5 \times 10^{-6}$ | 1.06 | E |
| Cardiac disorder | 10061024 | 1.31 [1.18,1.45] | $< 2.5 \times 10^{-3}$ | 1.06 | E |
| Hypotension | 10021097 | 1.18 [1.11,1.25] | $< 1.3 \times 10^{-4}$ | 1.05 | E |

| | | | | | |
|---|---|---|---|---|---|
| Urinary tract infection | 10046571 | 1.37 [1.29,1.46] | $< 2.3 \times 10^{-19}$ | 1.04 | E |
| Pain in extremity | 10033425 | 1.21 [1.15,1.28] | $< 1.4 \times 10^{-8}$ | 1.04 | E |
| Renal injury | 10061481 | 0.24 [0.20,0.29] | $< 6.8 \times 10^{-57}$ | 1.03 | P |
| Neuropathy peripheral | 10029331 | 1.92 [1.78,2.08] | $< 7.1 \times 10^{-60}$ | 1.02 | E |
| Cardiac arrest | 10007515 | 1.93 [1.76,2.11] | $< 1.2 \times 10^{-43}$ | 1.00 | E |
| Bladder cancer | 10005003 | 2.59 [1.96,3.43] | $< 2.2 \times 10^{-8}$ | 0.95 | E |
| Pyrexia | 10037660 | 1.20 [1.14,1.26] | $< 3.5 \times 10^{-8}$ | 0.93 | E |
| Tubulointerstitial nephritis | 10048302 | 0.61 [0.51,0.74] | $< 5.0 \times 10^{-3}$ | 0.92 | P |
| Fluid overload | 10016803 | 1.38 [1.21,1.58] | $< 2.3 \times 10^{-2}$ | 0.91 | E |
| Hypertension | 10020772 | 1.26 [1.18,1.36] | $< 1.8 \times 10^{-6}$ | 0.88 | E |
| Gait disturbance | 10017577 | 1.37 [1.27,1.47] | $< 4.5 \times 10^{-14}$ | 0.85 | E |
| Respiratory arrest | 10038669 | 6.85 [5.46,8.60] | $< 1.1 \times 10^{-87}$ | 0.83 | E |
| Chest pain | 10008479 | 1.18 [1.10,1.26] | $< 2.8 \times 10^{-2}$ | 0.74 | E |
| Visual acuity reduced | 10047531 | 2.76 [2.05,3.71] | $< 1.8 \times 10^{-8}$ | 0.71 | E |
| Pneumonia | 10035664 | 1.14 [1.09,1.19] | $< 2.2 \times 10^{-4}$ | 0.71 | E |
| Hypoacusis | 10048865 | 1.82 [1.60,2.08] | $< 7.8 \times 10^{-16}$ | 0.69 | E |
| End stage renal disease | 10077512 | 0.38 [0.33,0.44] | $< 1.6 \times 10^{-33}$ | 0.67 | P |
| Psoriatic arthropathy | 10037162 | 1.43 [1.28,1.59] | $< 3.1 \times 10^{-6}$ | 0.64 | E |
| Cognitive disorder | 10057668 | 1.46 [1.26,1.70] | $< 3.7 \times 10^{-3}$ | 0.62 | E |
| Rectal haemorrhage | 10038063 | 1.55 [1.33,1.81] | $< 1.8 \times 10^{-4}$ | 0.59 | E |
| Memory impairment | 10027175 | 1.35 [1.25,1.46] | $< 2.5 \times 10^{-10}$ | 0.59 | E |
| Neurotoxicity | 10029350 | 1.45 [1.25,1.69] | $< 1.1 \times 10^{-2}$ | 0.58 | E |
| Chest discomfort | 10008469 | 1.27 [1.16,1.40] | $< 2.4 \times 10^{-3}$ | 0.53 | E |
| Insomnia | 10022437 | 1.19 [1.12,1.26] | $< 2.1 \times 10^{-4}$ | 0.52 | E |
| Cough | 10011224 | 1.23 [1.17,1.30] | $< 7.3 \times 10^{-10}$ | 0.48 | E |
| Confusional state | 10010305 | 1.40 [1.30,1.51] | $< 6.4 \times 10^{-15}$ | 0.46 | E |
| Nausea | 10028813 | 0.93 [0.90,0.96] | $< 1.7 \times 10^{-2}$ | 0.41 | P |
| Gastrointestinal haemorrhage | 10017955 | 1.68 [1.54,1.82] | $< 2.2 \times 10^{-31}$ | 0.41 | E |
| Arthralgia | 10003239 | 1.20 [1.15,1.26] | $< 1.7 \times 10^{-10}$ | 0.39 | E |
| Osteonecrosis of jaw | 10064658 | 0.57 [0.45,0.72] | $< 1.3 \times 10^{-2}$ | 0.33 | P |
| Neoplasm progression | 10061309 | 1.61 [1.43,1.82] | $< 1.6 \times 10^{-11}$ | 0.27 | E |
| Anxiety | 10002855 | 1.25 [1.17,1.33] | $< 9.0 \times 10^{-8}$ | 0.18 | E |
| Dyspnoea | 10013968 | 1.19 [1.15,1.24] | $< 3.1 \times 10^{-16}$ | 0.15 | E |
| Agranulocytosis | 10001507 | 0.38 [0.27,0.52] | $< 4.3 \times 10^{-6}$ | 0.15 | P |
| Oedema peripheral | 10030124 | 1.50 [1.37,1.64] | $< 3.0 \times 10^{-15}$ | 0.15 | E |
| Fluid retention | 10016807 | 1.44 [1.32,1.58] | $< 8.1 \times 10^{-12}$ | 0.13 | E |
| Tremor | 10044565 | 1.20 [1.11,1.29] | $< 3.8 \times 10^{-2}$ | 0.06 | E |
| Haematuria | 10018867 | 1.43 [1.23,1.66] | $< 3.9 \times 10^{-2}$ | 0.02 | E |

**Table S2: Summary of adverse events identified by our approach in male patients.** Shown are 19 adverse events including 16 enriched and 3 purified during the pandemic, which are identified in male subpopulation. The $\beta$ represents the association between adverse event and the pandemic period (p-value is measured by Fisher's exact test, adjusted by Bonferroni correction). The adverse events are sorted in descending order of PAEAI. The 'E' and 'P' in the 'E\P' column denotes this adverse event is enriched or purified, respectively. More notations are the same as in SI Table S1. PEx: pulmonary exacerbations.

| Adverse event | MedDRA ID | $\beta$ (95% CI) | p-value | PAEAI (↓) | E/P |
|---|---|---|---|---|---|
| Hypogammaglobulinaemia | 10020983 | 22.32 [9.11,54.69] | $< 1.6 \times 10^{-23}$ | 1.91 | E |
| Alcoholism | 10001639 | 1.90 [1.45,2.49] | $< 1.7 \times 10^{-2}$ | 1.79 | E |
| Delusion | 10012239 | 2.96 [2.28,3.85] | $< 2.1 \times 10^{-14}$ | 1.59 | E |
| Neutrophilia | 10029379 | 4.20 [2.92,6.04] | $< 3.2 \times 10^{-14}$ | 1.48 | E |
| Dementia | 10012267 | 1.83 [1.41,2.38] | $< 2.8 \times 10^{-2}$ | 1.24 | E |
| Hallucination | 10019063 | 2.79 [2.46,3.18] | $< 4.9 \times 10^{-58}$ | 1.23 | E |
| Infective PEx of cystic fibrosis | 10070608 | 0.54 [0.44,0.67] | $< 3.9 \times 10^{-5}$ | 1.23 | P |
| Cardiac arrest | 10007515 | 1.89 [1.66,2.16] | $< 2.8 \times 10^{-18}$ | 1.12 | E |
| Respiratory failure | 10038695 | 1.40 [1.23,1.60] | $< 4.3 \times 10^{-3}$ | 0.96 | E |
| Neuropathy peripheral | 10029331 | 2.32 [2.05,2.63] | $< 1.0 \times 10^{-38}$ | 0.95 | E |
| Internal haemorrhage | 10075192 | 2.41 [1.70,3.42] | $< 2.4 \times 10^{-3}$ | 0.90 | E |
| Respiratory arrest | 10038669 | 8.19 [5.64,11.90] | $< 9.6 \times 10^{-41}$ | 0.80 | E |
| Headache | 10019211 | 0.83 [0.78,0.89] | $< 4.3 \times 10^{-4}$ | 0.76 | P |
| Bradycardia | 10006093 | 1.59 [1.32,1.93] | $< 1.0 \times 10^{-2}$ | 0.65 | E |
| Rectal haemorrhage | 10038063 | 1.86 [1.44,2.40] | $< 8.1 \times 10^{-3}$ | 0.61 | E |
| Gastrointestinal haemorrhage | 10017955 | 1.78 [1.58,2.02] | $< 3.8 \times 10^{-17}$ | 0.59 | E |
| Confusional state | 10010305 | 1.54 [1.38,1.73] | $< 4.0 \times 10^{-10}$ | 0.26 | E |
| Gynaecomastia | 10018800 | 0.55 [0.43,0.70] | $< 4.5 \times 10^{-3}$ | 0.21 | P |
| Cytokine release syndrome | 10052015 | 1.81 [1.45,2.24] | $< 3.7 \times 10^{-4}$ | 0.08 | E |

**Table S3: Summary of adverse events identified by our approach in female patients.** Shown are 44 adverse events including 38 enriched and 6 purified during the pandemic, identified in female subpopulation. The $\beta$ represents the association between adverse event and the pandemic period (p-value is measured by Fisher's exact test, adjusted by Bonferroni correction). The adverse events are sorted in descending order of PAEAI. The 'E' and 'P' in the 'E\P' column denotes this adverse event is enriched or purified, respectively. More notations are the same as in SI Table S1. PEx: pulmonary exacerbations, PPE: Palmar-plantar erythrodysaesthesia syndrome.

| Adverse event | MedDRA ID | $\beta$ (95% CI) | p-value | PAEAI (↓) | E/P |
|---|---|---|---|---|---|
| Premature delivery | 10036595 | 1.84 [1.50,2.25] | $< 2.4 \times 10^{-5}$ | 2.53 | E |
| Dizziness postural | 10013578 | 2.46 [1.76,3.45] | $< 4.7 \times 10^{-4}$ | 1.44 | E |
| Hypotension | 10021097 | 1.26 [1.17,1.37] | $< 6.0 \times 10^{-5}$ | 1.37 | E |
| Pulmonary hypertension | 10037400 | 1.73 [1.47,2.04] | $< 1.7 \times 10^{-7}$ | 1.34 | E |
| Hallucination | 10019063 | 2.10 [1.83,2.41] | $< 1.6 \times 10^{-23}$ | 1.26 | E |
| Neuropathy peripheral | 10029331 | 1.84 [1.66,2.05] | $< 7.4 \times 10^{-27}$ | 1.23 | E |
| Pain in extremity | 10033425 | 1.21 [1.14,1.29] | $< 1.9 \times 10^{-5}$ | 1.21 | E |
| Infective PEx of cystic fibrosis | 10070608 | 0.50 [0.42,0.60] | $< 3.6 \times 10^{-11}$ | 1.21 | P |
| Melanocytic naevus | 10027145 | 3.08 [2.15,4.41] | $< 4.0 \times 10^{-7}$ | 1.20 | E |
| Pain in jaw | 10033433 | 1.77 [1.57,2.00] | $< 3.0 \times 10^{-17}$ | 1.10 | E |
| Hypoxia | 10021143 | 1.51 [1.31,1.75] | $< 2.3 \times 10^{-4}$ | 1.03 | E |
| Urinary tract infection | 10046571 | 1.38 [1.28,1.48] | $< 1.0 \times 10^{-13}$ | 0.98 | E |
| Confusional state | 10010305 | 1.28 [1.15,1.42] | $< 4.4 \times 10^{-2}$ | 0.94 | E |
| Fluid overload | 10016803 | 1.56 [1.32,1.86] | $< 2.2 \times 10^{-3}$ | 0.92 | E |
| Cardiac arrest | 10007515 | 2.04 [1.78,2.34] | $< 9.1 \times 10^{-23}$ | 0.88 | E |
| Gait disturbance | 10017577 | 1.34 [1.22,1.46] | $< 4.4 \times 10^{-6}$ | 0.84 | E |
| Hypertension | 10020772 | 1.33 [1.21,1.46] | $< 6.0 \times 10^{-5}$ | 0.83 | E |
| Pneumonia | 10035664 | 1.27 [1.20,1.35] | $< 8.1 \times 10^{-11}$ | 0.83 | E |
| Insomnia | 10022437 | 1.21 [1.11,1.31] | $< 3.6 \times 10^{-2}$ | 0.83 | E |
| Pyrexia | 10037660 | 1.31 [1.22,1.41] | $< 1.5 \times 10^{-9}$ | 0.81 | E |
| Hypoacusis | 10048865 | 1.96 [1.64,2.33] | $< 1.0 \times 10^{-10}$ | 0.77 | E |
| Psoriatic arthropathy | 10037162 | 1.51 [1.32,1.73] | $< 8.2 \times 10^{-6}$ | 0.75 | E |
| Respiratory arrest | 10038669 | 9.40 [6.60,13.40] | $< 7.1 \times 10^{-55}$ | 0.74 | E |
| Influenza | 10022000 | 1.33 [1.20,1.46] | $< 7.0 \times 10^{-5}$ | 0.70 | E |
| Chest pain | 10008479 | 1.27 [1.17,1.39] | $< 2.6 \times 10^{-4}$ | 0.68 | E |
| Chest discomfort | 10008469 | 1.44 [1.29,1.62] | $< 1.7 \times 10^{-6}$ | 0.68 | E |
| Cough | 10011224 | 1.33 [1.24,1.42] | $< 2.7 \times 10^{-12}$ | 0.66 | E |
| Renal impairment | 10062237 | 1.53 [1.27,1.84] | $< 4.1 \times 10^{-2}$ | 0.59 | E |
| Gastrointestinal haemorrhage | 10017955 | 2.03 [1.78,2.31] | $< 2.7 \times 10^{-23}$ | 0.57 | E |
| Chronic kidney disease | 10064848 | 0.41 [0.34,0.48] | $< 4.7 \times 10^{-25}$ | 0.55 | P |
| PPE syndrome | 10033553 | 0.53 [0.40,0.69] | $< 1.7 \times 10^{-2}$ | 0.47 | P |

| | | | | | |
|---|---|---|---|---|---|
| Renal injury | 10061481 | 0.33 [0.25,0.44] | $< 1.2 \times 10^{-11}$ | 0.43 | P |
| Neoplasm progression | 10061309 | 1.75 [1.52,2.01] | $< 8.9 \times 10^{-12}$ | 0.42 | E |
| Palpitations | 10033557 | 1.28 [1.15,1.41] | $< 3.0 \times 10^{-2}$ | 0.41 | E |
| Anxiety | 10002855 | 1.34 [1.24,1.45] | $< 2.3 \times 10^{-9}$ | 0.39 | E |
| Back pain | 10003988 | 1.21 [1.12,1.30] | $< 3.2 \times 10^{-3}$ | 0.39 | E |
| Nausea | 10028813 | 0.90 [0.87,0.93] | $< 1.0 \times 10^{-4}$ | 0.20 | P |
| Dyspnoea | 10013968 | 1.25 [1.19,1.31] | $< 3.9 \times 10^{-16}$ | 0.19 | E |
| Fluid retention | 10016807 | 1.55 [1.39,1.74] | $< 5.1 \times 10^{-11}$ | 0.17 | E |
| Memory impairment | 10027175 | 1.34 [1.21,1.47] | $< 3.1 \times 10^{-5}$ | 0.12 | E |
| Flushing | 10016825 | 1.28 [1.15,1.42] | $< 2.4 \times 10^{-2}$ | 0.10 | E |
| Oedema peripheral | 10030124 | 1.60 [1.42,1.79] | $< 1.1 \times 10^{-11}$ | 0.07 | E |
| Arthralgia | 10003239 | 1.17 [1.11,1.25] | $< 6.4 \times 10^{-4}$ | 0.02 | E |
| Cardio-respiratory arrest | 10007617 | 0.41 [0.33,0.52] | $< 3.7 \times 10^{-11}$ | 0.01 | P |

**Table S4: Summary of adverse events identified by our approach in young patients.** Our framework detects two adverse events (one enriched and one purified), which have significant different reporting pattern in the pandemic compared to before the pandemic, in young patients. The $\beta$ represents the association between adverse event and the pandemic period (p-value is measured by Fisher's exact test, adjusted by Bonferroni correction). The adverse events are sorted in descending order of PAEAI. The 'E' and 'P' in the 'E\P' column denotes this adverse event is enriched or purified, respectively. More notations are the same as in SI Table S1. PEx: pulmonary exacerbations.

| Adverse event | MedDRA ID | $\beta$ (95% CI) | p-value | PAEAI (↓) | E/P |
|---|---|---|---|---|---|
| Infective PEx of cystic fibrosis | 10070608 | 0.48 [0.38,0.62] | $< 7.3 \times 10^{-6}$ | 1.33 | P |
| Pyrexia | 10037660 | 1.50 [1.26,1.78] | $< 1.1 \times 10^{-2}$ | 1.08 | E |

**Table S5: Summary of adverse events identified by our approach in adult patients.** Shown are 37 adverse events, including 35 enriched and 2 purified, identified in adults. The $\beta$ represents the association between adverse event and the pandemic period (p-value is measured by Fisher's exact test, adjusted by Bonferroni correction). The adverse events are sorted in descending order of PAEAI. The 'E' and 'P' in the 'E\P' column denotes this adverse event is enriched or purified, respectively. More notations are the same as in SI Table S1. PEx: pulmonary exacerbations.

| Adverse event | MedDRA ID | $\beta$ (95% CI) | p-value | PAEAI (↓) | E/P |
|---|---|---|---|---|---|
| Pain in jaw | 10033433 | 1.93 [1.65,2.25] | $< 1.1 \times 10^{-13}$ | 1.61 | E |
| Premature delivery | 10036595 | 2.32 [1.86,2.90] | $< 1.7 \times 10^{-10}$ | 1.58 | E |
| Pulmonary hypertension | 10037400 | 2.14 [1.71,2.69] | $< 1.7 \times 10^{-7}$ | 1.48 | E |
| Pulmonary arterial hypertension | 10064911 | 1.82 [1.48,2.25] | $< 1.0 \times 10^{-4}$ | 1.43 | E |
| Neutrophilia | 10029379 | 4.96 [3.53,6.96] | $< 3.8 \times 10^{-22}$ | 1.34 | E |
| Acute kidney injury | 10069339 | 1.41 [1.28,1.55] | $< 1.5 \times 10^{-8}$ | 1.32 | E |
| Melanocytic naevus | 10027145 | 3.64 [2.52,5.26] | $< 8.5 \times 10^{-10}$ | 1.32 | E |
| Respiratory failure | 10038695 | 1.49 [1.29,1.72] | $< 3.0 \times 10^{-4}$ | 1.29 | E |
| Pyrexia | 10037660 | 1.29 [1.19,1.40] | $< 9.1 \times 10^{-6}$ | 1.22 | E |
| Hypoxia | 10021143 | 1.58 [1.33,1.87] | $< 1.0 \times 10^{-3}$ | 1.14 | E |
| Pneumonia | 10035664 | 1.22 [1.13,1.33] | $< 1.3 \times 10^{-2}$ | 1.12 | E |
| Neuropathy peripheral | 10029331 | 1.84 [1.58,2.15] | $< 3.9 \times 10^{-11}$ | 1.10 | E |
| Haemorrhage | 10055798 | 1.93 [1.61,2.32] | $< 5.0 \times 10^{-9}$ | 1.07 | E |
| Cardiac arrest | 10007515 | 2.71 [2.37,3.09] | $< 3.0 \times 10^{-50}$ | 1.01 | E |
| Rectal haemorrhage | 10038063 | 1.99 [1.50,2.64] | $< 9.7 \times 10^{-3}$ | 0.99 | E |
| Hypotension | 10021097 | 1.59 [1.44,1.75] | $< 1.6 \times 10^{-17}$ | 0.88 | E |
| Bone marrow failure | 10065553 | 0.41 [0.29,0.59] | $< 4.9 \times 10^{-3}$ | 0.87 | P |
| Neutropenia | 10029354 | 1.36 [1.21,1.53] | $< 1.4 \times 10^{-3}$ | 0.87 | E |
| Pain in extremity | 10033425 | 1.30 [1.19,1.41] | $< 6.8 \times 10^{-6}$ | 0.85 | E |
| Respiratory arrest | 10038669 | 20.18 [13.26,30.72] | $< 1.2 \times 10^{-99}$ | 0.85 | E |
| Tachycardia | 10043071 | 1.46 [1.26,1.69] | $< 5.4 \times 10^{-3}$ | 0.84 | E |

| | | | | | |
|---|---|---|---|---|---|
| Chest pain | 10008479 | 1.32 [1.20,1.47] | $< 5.3 \times 10^{-4}$ | 0.78 | E |
| Gastrointestinal haemorrhage | 10017955 | 2.82 [2.35,3.38] | $< 3.0 \times 10^{-28}$ | 0.77 | E |
| Psoriatic arthropathy | 10037162 | 1.74 [1.48,2.04] | $< 6.8 \times 10^{-8}$ | 0.73 | E |
| Fluid overload | 10016803 | 1.67 [1.37,2.03] | $< 1.7 \times 10^{-3}$ | 0.73 | E |
| Arthralgia | 10003239 | 1.23 [1.14,1.33] | $< 5.5 \times 10^{-4}$ | 0.72 | E |
| Palpitations | 10033557 | 1.43 [1.25,1.63] | $< 1.1 \times 10^{-3}$ | 0.65 | E |
| Cough | 10011224 | 1.53 [1.39,1.67] | $< 5.7 \times 10^{-16}$ | 0.63 | E |
| Psoriasis | 10037153 | 1.35 [1.23,1.48] | $< 3.5 \times 10^{-7}$ | 0.62 | E |
| Cytokine release syndrome | 10052015 | 1.84 [1.47,2.30] | $< 4.6 \times 10^{-4}$ | 0.62 | E |
| Menorrhagia | 10027313 | 1.81 [1.43,2.30] | $< 5.8 \times 10^{-3}$ | 0.61 | E |
| Anxiety | 10002855 | 1.36 [1.23,1.50] | $< 1.7 \times 10^{-5}$ | 0.56 | E |
| Neoplasm progression | 10061309 | 1.87 [1.54,2.28] | $< 1.4 \times 10^{-6}$ | 0.37 | E |
| Dyspnoea | 10013968 | 1.31 [1.23,1.39] | $< 3.3 \times 10^{-14}$ | 0.36 | E |
| Chronic kidney disease | 10064848 | 0.58 [0.47,0.72] | $< 2.6 \times 10^{-3}$ | 0.35 | P |
| Oedema peripheral | 10030124 | 1.57 [1.35,1.83] | $< 4.5 \times 10^{-5}$ | 0.30 | E |
| Sinusitis | 10040753 | 1.52 [1.34,1.72] | $< 2.9 \times 10^{-7}$ | 0.03 | E |

**Table S6: Summary of adverse events identified by our approach in elderly patients.** Shown are 19 adverse events (18 enriched and 1 purified), identified in aged individuals. The $\beta$ represents the association between adverse event and the pandemic period (p-value is measured by Fisher's exact test, adjusted by Bonferroni correction). The adverse events are sorted in descending order of PAEAI. The 'E' and 'P' in the 'E\P' column denotes this adverse event is enriched or purified, respectively. More notations are the same as in SI Table S1.

| Adverse event | MedDRA ID | $\beta$ (95% CI) | p-value | PAEAI (↓) | E/P |
|---|---|---|---|---|---|
| Aggression | 10001488 | 2.42 [1.70,3.45] | $< 2.1 \times 10^{-3}$ | 1.46 | E |
| Vitreous floaters | 10047654 | 8.11 [4.97,13.23] | $< 1.5 \times 10^{-22}$ | 1.40 | E |
| Abnormal behaviour | 10061422 | 2.85 [2.03,3.98] | $< 4.9 \times 10^{-7}$ | 1.16 | E |
| Delusion | 10012239 | 2.79 [2.09,3.72] | $< 9.3 \times 10^{-10}$ | 1.07 | E |
| Haematuria | 10018867 | 1.98 [1.55,2.54] | $< 1.5 \times 10^{-4}$ | 1.04 | E |
| Hallucination | 10019063 | 2.58 [2.24,2.97] | $< 1.1 \times 10^{-39}$ | 1.01 | E |
| Cardiac arrest | 10007515 | 1.59 [1.34,1.89] | $< 7.5 \times 10^{-4}$ | 0.99 | E |
| Internal haemorrhage | 10075192 | 2.92 [2.01,4.24] | $< 1.2 \times 10^{-5}$ | 0.97 | E |
| Dementia | 10012267 | 1.97 [1.51,2.58] | $< 2.4 \times 10^{-3}$ | 0.94 | E |
| General physical health deterioration | 10049438 | 1.97 [1.50,2.59] | $< 4.8 \times 10^{-3}$ | 0.91 | E |
| Rectal haemorrhage | 10038063 | 1.88 [1.47,2.39] | $< 1.1 \times 10^{-3}$ | 0.90 | E |
| Urinary tract infection | 10046571 | 1.29 [1.16,1.43] | $< 1.7 \times 10^{-2}$ | 0.89 | E |
| Neuropathy peripheral | 10029331 | 1.58 [1.36,1.84] | $< 1.0 \times 10^{-5}$ | 0.83 | E |
| Acute kidney injury | 10069339 | 1.35 [1.21,1.50] | $< 1.4 \times 10^{-4}$ | 0.83 | E |
| Gastrointestinal haemorrhage | 10017955 | 2.00 [1.78,2.26] | $< 2.9 \times 10^{-28}$ | 0.75 | E |
| Hypoacusis | 10048865 | 2.18 [1.78,2.66] | $< 2.1 \times 10^{-11}$ | 0.73 | E |
| Confusional state | 10010305 | 1.51 [1.34,1.70] | $< 9.0 \times 10^{-8}$ | 0.73 | E |
| Nausea | 10028813 | 0.85 [0.80,0.91] | $< 2.4 \times 10^{-2}$ | 0.63 | P |
| Neoplasm progression | 10061309 | 1.92 [1.61,2.30] | $< 5.7 \times 10^{-10}$ | 0.47 | E |

**Table S7: Associations between adverse events and the pandemic.** The 'Occur 2019' and 'Nonoccur 2019' denote the number of submitted reports that contain and not contain the specific adverse event in 2019 (March 11 – September 30), respectively. Similarly, 'Occur 2020' and 'Nonoccur 2020' denote the statistics in the same period of 2020. On the basis of these numbers, we calculate the defined $\beta$ following Eq. 2 (Methods), which measures the association between the adverse event and the pandemic. The p-value is measured by Fisher's exact test followed by Bonferroni correction. In this table, we focus on the adverse events that involved in at least 100 reports either in 2019 or 2020, and present the top 100 adverse events with the lowest p-values. Then, the adverse events are sorted in descending order of $\beta$. Higher $\beta$ and smaller p-value indicate the adverse event has stronger association with the pandemic. PEx: pulmonary exacerbation; IEC: Immune effector cell.

| Adverse event | Occur 2019 | Nonoccur 2019 | Occur 2020 | Nonoccur 2020 | $\beta$ (95% CI) (↓) | p-value |
|---|---|---|---|---|---|---|
| Vitritis | 8 | 220,912 | 105 | 211,047 | 13.74 [6.69,28.19] | $< 8.0 \times 10^{-20}$ |
| Hypogammaglobulinaemia | 27 | 220,893 | 198 | 210,954 | 7.68 [5.14,11.48] | $< 4.0 \times 10^{-31}$ |
| Respiratory arrest | 86 | 220,834 | 562 | 210,590 | 6.85 [5.46,8.60] | $< 1.1 \times 10^{-87}$ |
| Vitreous floaters | 62 | 220,858 | 228 | 210,924 | 3.85 [2.91,5.10] | $< 3.0 \times 10^{-21}$ |
| Eye inflammation | 49 | 220,871 | 160 | 210,992 | 3.42 [2.48,4.71] | $< 3.1 \times 10^{-12}$ |
| Uveitis | 53 | 220,867 | 161 | 210,991 | 3.18 [2.33,4.34] | $< 4.3 \times 10^{-11}$ |
| Melanocytic naevus | 55 | 220,865 | 146 | 211,006 | 2.78 [2.04,3.79] | $< 8.7 \times 10^{-8}$ |
| Hypoalbuminaemia | 40 | 220,880 | 106 | 211,046 | 2.77 [1.93,3.99] | $< 6.3 \times 10^{-5}$ |
| Visual acuity reduced | 60 | 220,860 | 158 | 210,994 | 2.76 [2.05,3.71] | $< 1.8 \times 10^{-8}$ |
| Bladder cancer | 69 | 220,851 | 171 | 210,981 | 2.59 [1.96,3.43] | $< 2.2 \times 10^{-8}$ |
| Neutrophilia | 103 | 220,817 | 250 | 210,902 | 2.54 [2.02,3.20] | $< 7.6 \times 10^{-13}$ |
| Delusion | 143 | 220,777 | 345 | 210,807 | 2.53 [2.08,3.07] | $< 1.6 \times 10^{-18}$ |
| Neuroleptic malignant syndrome | 44 | 220,876 | 106 | 211,046 | 2.52 [1.77,3.58] | $< 7.2 \times 10^{-4}$ |
| Dystonia | 66 | 220,854 | 158 | 210,994 | 2.51 [1.88,3.34] | $< 7.0 \times 10^{-7}$ |
| Shock haemorrhagic | 44 | 220,876 | 105 | 211,047 | 2.50 [1.76,3.55] | $< 1.0 \times 10^{-3}$ |
| Internal haemorrhage | 86 | 220,834 | 203 | 210,949 | 2.47 [1.92,3.18] | $< 2.0 \times 10^{-9}$ |
| Haemorrhagic stroke | 61 | 220,859 | 142 | 211,010 | 2.44 [1.80,3.29] | $< 1.4 \times 10^{-5}$ |
| Hypothermia | 57 | 220,863 | 133 | 211,019 | 2.44 [1.79,3.33] | $< 4.0 \times 10^{-5}$ |
| Hallucination | 699 | 220,221 | 1,610 | 209,542 | 2.42 [2.21,2.65] | $< 5.7 \times 10^{-88}$ |
| Sensitivity to weather change | 48 | 220,872 | 102 | 211,050 | 2.22 [1.58,3.13] | $< 2.5 \times 10^{-2}$ |
| Dizziness postural | 97 | 220,823 | 190 | 210,962 | 2.05 [1.61,2.62] | $< 3.6 \times 10^{-5}$ |
| Tachypnoea | 91 | 220,829 | 171 | 210,981 | 1.97 [1.53,2.54] | $< 1.1 \times 10^{-3}$ |
| Eye pain | 251 | 220,669 | 473 | 210,679 | 1.97 [1.69,2.30] | $< 5.6 \times 10^{-15}$ |
| Lymphopenia | 95 | 220,825 | 178 | 210,974 | 1.96 [1.53,2.52] | $< 7.6 \times 10^{-4}$ |
| Cardiac arrest | 725 | 220,195 | 1,331 | 209,821 | 1.93 [1.76,2.11] | $< 1.2 \times 10^{-43}$ |
| Cytokine release syndrome | 282 | 220,638 | 520 | 210,632 | 1.93 [1.67,2.23] | $< 9.7 \times 10^{-16}$ |
| Neuropathy peripheral | 994 | 219,926 | 1,817 | 209,335 | 1.92 [1.78,2.08] | $< 7.1 \times 10^{-60}$ |
| Premature delivery | 154 | 220,766 | 267 | 210,885 | 1.82 [1.49,2.21] | $< 2.1 \times 10^{-5}$ |
| Hypoacusis | 350 | 220,570 | 609 | 210,543 | 1.82 [1.60,2.08] | $< 7.8 \times 10^{-16}$ |
| Pain in jaw | 562 | 220,358 | 934 | 210,218 | 1.74 [1.57,1.93] | $< 4.8 \times 10^{-22}$ |
| Dry eye | 246 | 220,674 | 400 | 210,752 | 1.70 [1.45,2.00] | $< 2.8 \times 10^{-7}$ |
| Encephalopathy | 262 | 220,658 | 422 | 210,730 | 1.69 [1.45,1.97] | $< 1.6 \times 10^{-7}$ |
| Lower limb fracture | 132 | 220,788 | 212 | 210,940 | 1.68 [1.35,2.09] | $< 2.3 \times 10^{-2}$ |
| Dementia | 206 | 220,714 | 331 | 210,821 | 1.68 [1.41,2.00] | $< 3.5 \times 10^{-5}$ |

| | | | | | | |
|---|---|---|---|---|---|---|
| Gastrointestinal haemorrhage | 914 | 220,006 | 1,462 | 209,690 | 1.68 [1.54,1.82] | $< 2.2 \times 10^{-31}$ |
| Restless legs syndrome | 159 | 220,761 | 251 | 210,901 | 1.65 [1.35,2.02] | $< 5.3 \times 10^{-3}$ |
| Ageusia | 211 | 220,709 | 329 | 210,823 | 1.63 [1.37,1.94] | $< 2.3 \times 10^{-4}$ |
| Neoplasm progression | 451 | 220,469 | 694 | 210,458 | 1.61 [1.43,1.82] | $< 1.6 \times 10^{-11}$ |
| Alcoholism | 177 | 220,743 | 267 | 210,885 | 1.58 [1.31,1.91] | $< 2.3 \times 10^{-2}$ |
| Rectal haemorrhage | 270 | 220,650 | 401 | 210,751 | 1.55 [1.33,1.81] | $< 1.8 \times 10^{-4}$ |
| Pulmonary hypertension | 384 | 220,536 | 553 | 210,599 | 1.51 [1.32,1.72] | $< 5.5 \times 10^{-6}$ |
| Nasal congestion | 570 | 220,350 | 823 | 210,329 | 1.51 [1.36,1.68] | $< 2.0 \times 10^{-10}$ |
| Oedema peripheral | 815 | 220,105 | 1,168 | 209,984 | 1.50 [1.37,1.64] | $< 3.0 \times 10^{-15}$ |
| Hypoxia | 574 | 220,346 | 809 | 210,343 | 1.48 [1.33,1.64] | $< 7.9 \times 10^{-9}$ |
| Rheumatoid arthritis | 526 | 220,394 | 736 | 210,416 | 1.47 [1.31,1.64] | $< 1.7 \times 10^{-7}$ |
| Cognitive disorder | 301 | 220,619 | 421 | 210,731 | 1.46 [1.26,1.70] | $< 3.7 \times 10^{-3}$ |
| Neurotoxicity | 293 | 220,627 | 406 | 210,746 | 1.45 [1.25,1.69] | $< 1.1 \times 10^{-2}$ |
| Fluid retention | 826 | 220,094 | 1,138 | 210,014 | 1.44 [1.32,1.58] | $< 8.0 \times 10^{-12}$ |
| Haematuria | 293 | 220,627 | 399 | 210,753 | 1.43 [1.23,1.66] | $< 3.9 \times 10^{-2}$ |
| Psoriatic arthropathy | 541 | 220,379 | 737 | 210,415 | 1.43 [1.28,1.59] | $< 3.1 \times 10^{-6}$ |
| Neoplasm malignant | 344 | 220,576 | 459 | 210,693 | 1.40 [1.21,1.61] | $< 2.5 \times 10^{-2}$ |
| Confusional state | 1,229 | 219,691 | 1,638 | 209,514 | 1.40 [1.30,1.51] | $< 6.3 \times 10^{-15}$ |
| Sinusitis | 1,048 | 219,872 | 1,388 | 209,764 | 1.39 [1.28,1.50] | $< 1.0 \times 10^{-11}$ |
| Bradycardia | 455 | 220,465 | 601 | 210,551 | 1.38 [1.22,1.56] | $< 1.7 \times 10^{-3}$ |
| Fluid overload | 376 | 220,544 | 496 | 210,656 | 1.38 [1.21,1.58] | $< 2.3 \times 10^{-2}$ |
| Urinary tract infection | 1,735 | 219,185 | 2,272 | 208,880 | 1.37 [1.29,1.46] | $< 2.3 \times 10^{-19}$ |
| Gait disturbance | 1,347 | 219,573 | 1,758 | 209,394 | 1.37 [1.27,1.47] | $< 4.5 \times 10^{-14}$ |
| Respiratory failure | 871 | 220,049 | 1,130 | 210,022 | 1.36 [1.24,1.49] | $< 1.0 \times 10^{-7}$ |
| Memory impairment | 1,152 | 219,768 | 1,482 | 209,670 | 1.35 [1.25,1.46] | $< 2.5 \times 10^{-10}$ |
| Epistaxis | 637 | 220,283 | 800 | 210,352 | 1.32 [1.18,1.46] | $< 2.4 \times 10^{-3}$ |
| Cardiac disorder | 668 | 220,252 | 834 | 210,318 | 1.31 [1.18,1.45] | $< 2.5 \times 10^{-3}$ |
| Chest discomfort | 826 | 220,094 | 1,005 | 210,147 | 1.27 [1.16,1.40] | $< 2.4 \times 10^{-3}$ |
| Hypertension | 1,358 | 219,562 | 1,638 | 209,514 | 1.26 [1.18,1.36] | $< 1.8 \times 10^{-6}$ |
| Anxiety | 1,777 | 219,143 | 2,113 | 209,039 | 1.25 [1.17,1.33] | $< 9.0 \times 10^{-8}$ |
| Peripheral swelling | 2,136 | 218,784 | 2,511 | 208,641 | 1.23 [1.16,1.31] | $< 1.5 \times 10^{-8}$ |
| Palpitations | 914 | 220,006 | 1,073 | 210,079 | 1.23 [1.13,1.34] | $< 4.8 \times 10^{-2}$ |
| Cough | 2,451 | 218,469 | 2,874 | 208,278 | 1.23 [1.17,1.30] | $< 7.3 \times 10^{-10}$ |
| Pain in extremity | 2,613 | 218,307 | 3,014 | 208,138 | 1.21 [1.15,1.28] | $< 1.4 \times 10^{-8}$ |
| Arthralgia | 3,266 | 217,654 | 3,744 | 207,408 | 1.20 [1.15,1.26] | $< 1.7 \times 10^{-10}$ |
| Pyrexia | 2,762 | 218,158 | 3,160 | 207,992 | 1.20 [1.14,1.26] | $< 3.5 \times 10^{-8}$ |
| Tremor | 1,260 | 219,660 | 1,439 | 209,713 | 1.20 [1.11,1.29] | $< 3.8 \times 10^{-2}$ |
| Insomnia | 1,971 | 218,949 | 2,238 | 208,914 | 1.19 [1.12,1.26] | $< 2.1 \times 10^{-4}$ |
| Dyspnoea | 5,414 | 215,506 | 6,130 | 205,022 | 1.19 [1.15,1.24] | $< 3.0 \times 10^{-16}$ |
| Chest pain | 1,595 | 219,325 | 1,790 | 209,362 | 1.18 [1.10,1.26] | $< 2.8 \times 10^{-2}$ |
| Hypotension | 2,286 | 218,634 | 2,570 | 208,582 | 1.18 [1.11,1.25] | $< 1.3 \times 10^{-4}$ |
| Back pain | 2,121 | 218,799 | 2,322 | 208,830 | 1.15 [1.08,1.22] | $< 5.5 \times 10^{-2}$ |
| Pneumonia | 3,708 | 217,212 | 4,021 | 207,131 | 1.14 [1.09,1.19] | $< 2.2 \times 10^{-4}$ |
| Nausea | 9,373 | 211,547 | 8,349 | 202,803 | 0.93 [0.90,0.96] | $< 1.7 \times 10^{-2}$ |
| Influenza like illness | 835 | 220,085 | 619 | 210,533 | 0.77 [0.70,0.86] | $< 1.4 \times 10^{-2}$ |
| Musculoskeletal pain | 562 | 220,358 | 400 | 210,752 | 0.74 [0.65,0.85] | $< 5.9 \times 10^{-2}$ |

| | | | | | | |
|---|---|---|---|---|---|---|
| Anaphylactic reaction | 653 | 220,267 | 459 | 210,693 | 0.73 [0.65,0.83] | $< 3.6 \times 10^{-3}$ |
| Tubulointerstitial nephritis | 278 | 220,642 | 163 | 210,989 | 0.61 [0.51,0.74] | $< 5.0 \times 10^{-3}$ |
| Haemorrhoids | 249 | 220,671 | 141 | 211,011 | 0.59 [0.48,0.73] | $< 4.6 \times 10^{-3}$ |
| Osteonecrosis of jaw | 198 | 220,722 | 107 | 211,045 | 0.57 [0.45,0.72] | $< 1.3 \times 10^{-2}$ |
| Photosensitivity reaction | 217 | 220,703 | 114 | 211,038 | 0.55 [0.44,0.69] | $< 1.4 \times 10^{-3}$ |
| Dermatitis allergic | 142 | 220,778 | 71 | 211,081 | 0.52 [0.39,0.70] | $< 5.2 \times 10^{-2}$ |
| Infective PEx of cystic fibrosis | 661 | 220,259 | 329 | 210,823 | 0.52 [0.46,0.59] | $< 2.5 \times 10^{-19}$ |
| Diverticulum | 120 | 220,800 | 55 | 211,097 | 0.48 [0.35,0.66] | $< 3.2 \times 10^{-2}$ |
| Hiatus hernia | 185 | 220,735 | 83 | 211,069 | 0.47 [0.36,0.61] | $< 3.1 \times 10^{-5}$ |
| Cardio-respiratory arrest | 473 | 220,447 | 212 | 210,940 | 0.47 [0.40,0.55] | $< 1.9 \times 10^{-17}$ |
| Bone marrow failure | 243 | 220,677 | 100 | 211,052 | 0.43 [0.34,0.54] | $< 1.4 \times 10^{-9}$ |
| End stage renal disease | 585 | 220,335 | 213 | 210,939 | 0.38 [0.33,0.44] | $< 1.6 \times 10^{-33}$ |
| Agranulocytosis | 141 | 220,779 | 51 | 211,101 | 0.38 [0.27,0.52] | $< 4.2 \times 10^{-6}$ |
| Chronic kidney disease | 2,113 | 218,807 | 640 | 210,512 | 0.31 [0.29,0.34] | $< 2.3 \times 10^{-165}$ |
| Large intestine polyp | 135 | 220,785 | 38 | 211,114 | 0.29 [0.21,0.42] | $< 4.0 \times 10^{-9}$ |
| Diverticulum intestinal | 109 | 220,811 | 27 | 211,125 | 0.26 [0.17,0.39] | $< 3.0 \times 10^{-8}$ |
| Renal injury | 556 | 220,364 | 127 | 211,025 | 0.24 [0.20,0.29] | $< 6.8 \times 10^{-57}$ |
| Coeliac disease | 143 | 220,777 | 29 | 211,123 | 0.21 [0.14,0.32] | $< 2.6 \times 10^{-14}$ |
| Granulocytopenia | 148 | 220,772 | 27 | 211,125 | 0.19 [0.13,0.29] | $< 2.4 \times 10^{-16}$ |
| Sprue-like enteropathy | 353 | 220,567 | 0 | 211,152 | 0.00 [0.00,0.00] | $< 2.4 \times 10^{-99}$ |

**Table S8: Associations between adverse events and drugs.** This table shows the number of patients who: exposed to the drug and have the adverse event (denoted by 'A'); exposed to the drug but not have the adverse event (denoted by 'B'); not exposed to the drug but have the adverse event (denoted by 'C'); not exposed to the drug and not have the adverse event (denoted by 'D'). Please note all the numbers (A, B, C, and D) are measured during the pandemic period (March 11 – September 30, 2020). We conduct patient matching and split patients into test group and control group. The size of control group is ten times of that of test group, which means that $10(A + B) = C + D$ (Methods). On the top of the statistics, we qualify the defined $\gamma$ following Eq. 4 (Methods), where 'A', 'B', 'C', and 'D' denote the measurement of $f(d_j, s_h, 2020)$, $f(d_j, \neg s_h, 2020)$, $f(\neg d_j, s_h, 2020)$, and $f(\neg d_j, \neg s_h, 2020)$, respectively. The p-value is measured by Fisher's exact test followed by Bonferroni correction. In this table, we provide the top 100 drug-adverse event pairs with the lowest p-values and sort them in descending order of $\gamma$. Higher $\gamma$ and smaller p-value indicate the adverse event has stronger association with the drug. The '- -' denotes infinity since the denominator is zero in Eq. 4. PEx: pulmonary exacerbation; IEC: Immune effector cell.

| Adverse event | Drug name | A | B | C | D | $\gamma$ (95% CI) (↓) | p-value |
|---|---|---|---|---|---|---|---|
| Bladder cancer | Pioglitazone | 104 | 245 | 1 | 3,489 | 1,481.04 [205.79,10,659.00] | $< 5.4 \times 10^{-111}$ |
| Vascular calcification | Warfarin | 75 | 2,863 | 1 | 29,379 | 769.62 [106.97,5,537.48] | $< 1.7 \times 10^{-74}$ |
| Delusion | Pimavanserin | 269 | 4,212 | 4 | 44,806 | 715.39 [266.42,1,920.97] | $< 4.9 \times 10^{-273}$ |
| Epstein-barr viraemia | Tacrolimus | 67 | 2,512 | 1 | 25,789 | 687.84 [95.45,4,957.02] | $< 3.5 \times 10^{-66}$ |
| Melanocytic naevus | Fingolimod | 118 | 1,258 | 2 | 13,758 | 645.25 [159.30,2,613.52] | $< 5.3 \times 10^{-119}$ |
| Hypogammaglobulinaemia | Fludarabine | 148 | 578 | 3 | 7,257 | 619.40 [196.88,1,948.67] | $< 9.9 \times 10^{-153}$ |
| Type I hypersensitivity | Famotidine | 55 | 2,376 | 1 | 24,309 | 562.71 [77.83,4,068.17] | $< 1.1 \times 10^{-53}$ |
| Haemorrhagic stroke | Rivaroxaban | 99 | 4,546 | 2 | 46,448 | 505.76 [124.69,2,051.34] | $< 9.7 \times 10^{-98}$ |
| Hypothermia | Risperidone | 47 | 814 | 1 | 8,609 | 497.08 [68.49,3,607.59] | $< 1.1 \times 10^{-45}$ |
| Diverticulum intestinal haemorrhagic | Rivaroxaban | 49 | 4,596 | 1 | 46,449 | 495.21 [68.36,3,587.17] | $< 2.5 \times 10^{-47}$ |
| Haematotoxicity | Vincristine | 55 | 1,076 | 2 | 11,308 | 289.01 [70.40,1,186.47] | $< 1.5 \times 10^{-52}$ |
| Hypogammaglobulinaemia | Cyclophosphamide | 153 | 2,024 | 6 | 21,764 | 274.20 [121.13,620.69] | $< 2.8 \times 10^{-149}$ |
| Type I hypersensitivity | Cetirizine | 56 | 2,100 | 3 | 21,557 | 191.62 [59.93,612.71] | $< 4.6 \times 10^{-52}$ |
| Hallucination | Istradefylline | 78 | 807 | 7 | 8,843 | 122.10 [56.16,265.48] | $< 4.9 \times 10^{-71}$ |
| Neurotoxicity | Tisagenlecleucel | 104 | 391 | 12 | 4,938 | 109.45 [59.69,200.71] | $< 2.4 \times 10^{-95}$ |
| Pain in jaw | Epoprostenol | 378 | 621 | 142 | 9,848 | 42.21 [34.24,52.04] | $< 3.1 \times 10^{-297}$ |
| Neoplasm progression | Axitinib | 64 | 426 | 18 | 4,882 | 40.75 [23.93,69.38] | $< 4.4 \times 10^{-49}$ |
| Neurotoxicity | Fludarabine | 107 | 619 | 35 | 7,225 | 35.68 [24.15,52.72] | $< 1.9 \times 10^{-80}$ |
| Pyrexia | Tisagenlecleucel | 201 | 294 | 101 | 4,849 | 32.82 [25.15,42.85] | $< 1.7 \times 10^{-146}$ |
| Haematuria | Rivaroxaban | 181 | 4,464 | 71 | 46,379 | 26.49 [20.09,34.91] | $< 9.6 \times 10^{-127}$ |
| Hallucination | Rasagiline | 69 | 213 | 42 | 2,778 | 21.43 [14.25,32.23] | $< 8.9 \times 10^{-44}$ |
| Hallucination | Carbidopa | 404 | 1,569 | 244 | 19,486 | 20.56 [17.40,24.30] | $< 1.9 \times 10^{-259}$ |
| Delusion | Carbidopa | 102 | 1,871 | 53 | 19,677 | 20.24 [14.47,28.30] | $< 4.0 \times 10^{-65}$ |
| Pulmonary hypertension | Macitentan | 295 | 4,421 | 167 | 46,993 | 18.78 [15.49,22.76] | $< 2.1 \times 10^{-185}$ |
| Confusional state | Pimavanserin | 367 | 4,114 | 214 | 44,596 | 18.59 [15.66,22.07] | $< 1.0 \times 10^{-229}$ |
| Rectal haemorrhage | Rivaroxaban | 178 | 4,467 | 107 | 46,343 | 17.26 [13.55,21.98] | $< 3.2 \times 10^{-108}$ |
| Internal haemorrhage | Rivaroxaban | 101 | 4,544 | 60 | 46,390 | 17.19 [12.47,23.69] | $< 6.9 \times 10^{-61}$ |
| Neurotoxicity | Cyclophosphamide | 112 | 2,065 | 70 | 21,700 | 16.81 [12.43,22.74] | $< 6.4 \times 10^{-67}$ |
| Dementia | Pimavanserin | 103 | 4,378 | 65 | 44,745 | 16.20 [11.85,22.13] | $< 1.0 \times 10^{-60}$ |
| Pyrexia | Fludarabine | 207 | 519 | 175 | 7,085 | 16.15 [12.96,20.12] | $< 1.1 \times 10^{-118}$ |
| Fluid retention | Macitentan | 513 | 4,203 | 457 | 46,703 | 12.47 [10.95,14.20] | $< 8.0 \times 10^{-271}$ |

| | | | | | | | |
|---|---|---|---|---|---|---|---|
| Hypotension | Tisagenlecleucel | 87 | 408 | 87 | 4,863 | 11.92 [8.71,16.31] | $< 1.8 \times 10^{-43}$ |
| Bradycardia | Remdesivir | 125 | 2,894 | 116 | 30,074 | 11.20 [8.67,14.46] | $< 1.9 \times 10^{-62}$ |
| Hallucination | Quetiapine | 163 | 1,553 | 161 | 16,999 | 11.08 [8.86,13.86] | $< 1.3 \times 10^{-80}$ |
| Pain in jaw | Selexipag | 261 | 1,617 | 278 | 18,502 | 10.74 [9.01,12.82] | $< 1.6 \times 10^{-126}$ |
| Pulmonary hypertension | Sildenafil | 174 | 2,661 | 171 | 28,179 | 10.78 [8.69,13.36] | $< 2.3 \times 10^{-85}$ |
| Oedema peripheral | Macitentan | 375 | 4,341 | 393 | 46,767 | 10.28 [8.89,11.88] | $< 6.6 \times 10^{-180}$ |
| Hypotension | Riociguat | 252 | 1,273 | 324 | 14,926 | 9.12 [7.66,10.86] | $< 2.1 \times 10^{-110}$ |
| Neuropathy peripheral | Bortezomib | 172 | 1,402 | 215 | 15,525 | 8.86 [7.20,10.91] | $< 1.0 \times 10^{-74}$ |
| Fluid retention | Epoprostenol | 150 | 849 | 199 | 9,791 | 8.69 [6.95,10.87] | $< 1.3 \times 10^{-63}$ |
| Dyspnoea | Oxygen | 284 | 484 | 522 | 7,158 | 8.05 [6.78,9.55] | $< 2.2 \times 10^{-106}$ |
| Oedema peripheral | Riociguat | 148 | 1,377 | 205 | 15,045 | 7.89 [6.34,9.81] | $< 1.0 \times 10^{-59}$ |
| Fluid overload | Sildenafil | 132 | 2,703 | 176 | 28,174 | 7.82 [6.22,9.83] | $< 1.8 \times 10^{-53}$ |
| Confusional state | Carbidopa | 144 | 1,829 | 202 | 19,528 | 7.61 [6.11,9.48] | $< 5.2 \times 10^{-57}$ |
| Fluid overload | Macitentan | 191 | 4,525 | 263 | 46,897 | 7.53 [6.23,9.09] | $< 5.6 \times 10^{-76}$ |
| Dyspnoea | Epoprostenol | 405 | 594 | 846 | 9,144 | 7.37 [6.38,8.52] | $< 1.3 \times 10^{-139}$ |
| Hypotension | Sacubitril | 148 | 1,396 | 226 | 15,214 | 7.14 [5.76,8.84] | $< 1.0 \times 10^{-55}$ |
| Pain in jaw | Tadalafil | 218 | 1,784 | 340 | 19,680 | 7.07 [5.93,8.44] | $< 1.0 \times 10^{-81}$ |
| Fluid retention | Riociguat | 152 | 1,373 | 236 | 15,014 | 7.04 [5.70,8.70] | $< 1.2 \times 10^{-56}$ |
| Respiratory failure | Remdesivir | 119 | 2,900 | 176 | 30,014 | 7.00 [5.53,8.86] | $< 1.3 \times 10^{-44}$ |
| Pain in jaw | Sildenafil | 255 | 2,580 | 396 | 27,954 | 6.98 [5.93,8.21] | $< 2.7 \times 10^{-95}$ |
| Chest pain | Epoprostenol | 171 | 828 | 287 | 9,703 | 6.98 [5.70,8.55] | $< 4.7 \times 10^{-62}$ |
| Cardiac arrest | Remdesivir | 128 | 2,891 | 191 | 29,999 | 6.95 [5.54,8.73] | $< 8.4 \times 10^{-48}$ |
| Pyrexia | Cyclophosphamide | 258 | 1,919 | 421 | 21,349 | 6.82 [5.80,8.02] | $< 5.0 \times 10^{-94}$ |
| Dyspnoea | Treprostinil | 789 | 2,041 | 1,520 | 26,780 | 6.81 [6.18,7.51] | $< 5.8 \times 10^{-273}$ |
| Fluid retention | Sildenafil | 236 | 2,599 | 380 | 27,970 | 6.68 [5.65,7.90] | $< 1.5 \times 10^{-85}$ |
| Pain in jaw | Macitentan | 379 | 4,337 | 622 | 46,538 | 6.54 [5.73,7.46] | $< 9.4 \times 10^{-136}$ |
| Hypotension | Macitentan | 450 | 4,266 | 755 | 46,405 | 6.48 [5.75,7.32] | $< 1.2 \times 10^{-159}$ |
| Dyspnoea | Selexipag | 576 | 1,302 | 1,212 | 17,568 | 6.41 [5.72,7.19] | $< 3.7 \times 10^{-187}$ |
| Pain in jaw | Riociguat | 152 | 1,373 | 263 | 14,987 | 6.31 [5.13,7.76] | $< 4.5 \times 10^{-52}$ |
| Gait disturbance | Pimavanserin | 168 | 4,313 | 290 | 44,520 | 5.98 [4.93,7.25] | $< 2.8 \times 10^{-56}$ |
| Fluid retention | Tadalafil | 196 | 1,806 | 360 | 19,660 | 5.93 [4.95,7.10] | $< 3.7 \times 10^{-64}$ |
| Chest pain | Macitentan | 375 | 4,341 | 678 | 46,482 | 5.92 [5.20,6.74] | $< 2.9 \times 10^{-124}$ |
| Fluid retention | Selexipag | 165 | 1,713 | 302 | 18,478 | 5.89 [4.84,7.17] | $< 9.1 \times 10^{-54}$ |
| Dyspnoea | Sildenafil | 753 | 2,082 | 1,639 | 26,711 | 5.89 [5.35,6.50] | $< 2.2 \times 10^{-232}$ |
| Dyspnoea | Tadalafil | 569 | 1,433 | 1,293 | 18,727 | 5.75 [5.14,6.43] | $< 4.4 \times 10^{-170}$ |
| Neuropathy peripheral | Acyclovir | 247 | 4,094 | 458 | 42,952 | 5.66 [4.83,6.63] | $< 4.9 \times 10^{-79}$ |
| Pain in extremity | Epoprostenol | 187 | 812 | 391 | 9,599 | 5.65 [4.68,6.83] | $< 7.4 \times 10^{-57}$ |
| Hypotension | Selexipag | 209 | 1,669 | 412 | 18,368 | 5.58 [4.69,6.64] | $< 8.7 \times 10^{-65}$ |
| Pain in jaw | Treprostinil | 216 | 2,614 | 415 | 27,885 | 5.55 [4.69,6.58] | $< 1.8 \times 10^{-67}$ |
| Cough | Treprostinil | 314 | 2,516 | 633 | 27,667 | 5.45 [4.74,6.28] | $< 7.4 \times 10^{-96}$ |
| Oedema peripheral | Sildenafil | 166 | 2,669 | 321 | 28,029 | 5.43 [4.48,6.58] | $< 6.5 \times 10^{-51}$ |
| Cough | Macitentan | 524 | 4,192 | 1,103 | 46,057 | 5.22 [4.68,5.82] | $< 2.6 \times 10^{-154}$ |
| Chest discomfort | Macitentan | 200 | 4,516 | 402 | 46,758 | 5.15 [4.34,6.12] | $< 5.2 \times 10^{-59}$ |
| Dyspnoea | Riociguat | 419 | 1,106 | 1,045 | 14,205 | 5.15 [4.53,5.86] | $< 2.9 \times 10^{-113}$ |
| Neuropathy peripheral | Dexamethasone | 358 | 6,844 | 746 | 71,274 | 5.00 [4.40,5.68] | $< 3.3 \times 10^{-103}$ |
| Pain in extremity | Selexipag | 245 | 1,633 | 557 | 18,223 | 4.91 [4.19,5.75] | $< 4.1 \times 10^{-67}$ |

| | | | | | | | |
|---|---|---|---|---|---|---|---|
| Chest pain | Sildenafil | 210 | 2,625 | 473 | 27,877 | 4.71 [3.99,5.57] | $< 2.3 \times 10^{-56}$ |
| Arthralgia | Selexipag | 230 | 1,648 | 556 | 18,224 | 4.57 [3.89,5.38] | $< 7.9 \times 10^{-59}$ |
| Neuropathy peripheral | Pomalidomide | 195 | 4,150 | 462 | 42,988 | 4.37 [3.69,5.18] | $< 6.6 \times 10^{-49}$ |
| Cough | Tadalafil | 213 | 1,789 | 557 | 19,463 | 4.16 [3.53,4.91] | $< 1.9 \times 10^{-49}$ |
| Pneumonia | Macitentan | 541 | 4,175 | 1,614 | 45,546 | 3.66 [3.30,4.05] | $< 1.1 \times 10^{-108}$ |
| Cough | Sildenafil | 242 | 2,593 | 747 | 27,603 | 3.45 [2.97,4.01] | $< 3.3 \times 10^{-45}$ |
| Pain in extremity | Macitentan | 424 | 4,292 | 1,378 | 45,782 | 3.28 [2.93,3.68] | $< 1.5 \times 10^{-74}$ |
| Type I hypersensitivity | Oxaliplatin | 70 | 456 | 0 | 5,260 | - - | $< 1.1 \times 10^{-72}$ |
| Haematotoxicity | Methotrexate | 44 | 4,682 | 0 | 47,260 | - - | $< 9.4 \times 10^{-44}$ |
| Diffuse large b-cell lymphoma recurrent | Tisagenlecleucel | 44 | 451 | 0 | 4,950 | - - | $< 1.8 \times 10^{-44}$ |
| Haematotoxicity | Carboplatin | 46 | 862 | 0 | 9,080 | - - | $< 3.2 \times 10^{-46}$ |
| Haematotoxicity | Thiotepa | 46 | 113 | 0 | 1,590 | - - | $< 1.2 \times 10^{-48}$ |
| Hypogammaglobulinaemia | Tisagenlecleucel | 151 | 344 | 0 | 4,950 | - - | $< 2.5 \times 10^{-165}$ |
| Neutrophilia | Clozapine | 238 | 1,756 | 0 | 19,940 | - - | $< 1.5 \times 10^{-251}$ |
| Haematotoxicity | Cisplatin | 53 | 531 | 0 | 5,840 | - - | $< 5.2 \times 10^{-54}$ |
| Haematotoxicity | Cyclophosphamide | 55 | 2,122 | 0 | 21,770 | - - | $< 2.1 \times 10^{-55}$ |
| Haematotoxicity | Etoposide | 55 | 705 | 0 | 7,600 | - - | $< 6.4 \times 10^{-56}$ |

**Table S9: Associations between drug-adverse event pairs and the pandemic.** The 'Occur 2019' and 'Nonoccur 2019' denote the number of submitted reports that contain and not contain the specific drug-adverse event pair in 2019 (March 11 – September 30), respectively. Similarly, 'Occur 2020' and 'Nonoccur 2020' denote the statistics in the same period of 2020. On the basis of these numbers, we calculate the defined $\delta$ following Eq. 5 (Methods). The p-value is measured by Fisher's exact test followed by Bonferroni correction. We provide the top 100 drug-adverse event pairs with the lowest p-values. Then, the pairs are sorted in descending order of $\delta$. Higher $\delta$ and smaller p-value indicate the pair has stronger connection with the pandemic. The '- -' denotes infinity since the denominator is zero in Eq. 5. PEx: pulmonary exacerbation; IEC: Immune effector cell.

| Adverse event | Drug name | Occur 2019 | Nonoccur 2019 | Occur 2020 | Nonoccur 2020 | $\delta$ (95% CI) (↓) | p-value |
|---|---|---|---|---|---|---|---|
| Hypogammaglobulinaemia | Tisagenlecleucel | 1 | 220,919 | 151 | 211,001 | 158.10 [22.13,1,129.69] | $< 5.2 \times 10^{-41}$ |
| Cough | Elexacaftor | 1 | 220,919 | 120 | 211,032 | 125.62 [17.55,899.14] | $< 1.8 \times 10^{-31}$ |
| Respiratory arrest | Alprazolam | 1 | 220,919 | 98 | 211,054 | 102.58 [14.31,735.56] | $< 1.9 \times 10^{-24}$ |
| Vascular calcification | Warfarin | 1 | 220,919 | 75 | 211,077 | 78.50 [10.91,564.59] | $< 1.5 \times 10^{-17}$ |
| Hypogammaglobulinaemia | Fludarabine | 2 | 220,918 | 149 | 211,003 | 78.00 [19.33,314.80] | $< 9.0 \times 10^{-39}$ |
| Hypoxia | Fludarabine | 1 | 220,919 | 60 | 211,092 | 62.79 [8.70,453.10] | $< 5.3 \times 10^{-13}$ |
| Haematotoxicity | Etoposide | 1 | 220,919 | 55 | 211,097 | 57.56 [7.97,415.95] | $< 1.7 \times 10^{-11}$ |
| Bladder cancer | Pioglitazone | 2 | 220,918 | 104 | 211,048 | 54.43 [13.43,220.56] | $< 4.2 \times 10^{-25}$ |
| Hypogammaglobulinaemia | Cyclophosphamide | 4 | 220,916 | 154 | 210,998 | 40.31 [14.94,108.77] | $< 2.0 \times 10^{-37}$ |
| Haematotoxicity | Vincristine | 2 | 220,918 | 55 | 211,097 | 28.78 [7.02,117.99] | $< 2.9 \times 10^{-10}$ |
| Respiratory arrest | Fentanyl | 3 | 220,917 | 73 | 211,079 | 25.47 [8.03,80.81] | $< 1.2 \times 10^{-14}$ |
| Respiratory arrest | Oxycodone | 3 | 220,917 | 62 | 211,090 | 21.63 [6.79,68.90] | $< 3.3 \times 10^{-11}$ |
| Pain in extremity | Certolizumab pegol | 4 | 220,916 | 77 | 211,075 | 20.15 [7.37,55.05] | $< 8.8 \times 10^{-15}$ |
| Dyspnoea | Certolizumab pegol | 4 | 220,916 | 63 | 211,089 | 16.48 [6.00,45.29] | $< 1.7 \times 10^{-10}$ |
| Respiratory arrest | Acetaminophen | 8 | 220,912 | 114 | 211,038 | 14.92 [7.28,30.55] | $< 1.6 \times 10^{-21}$ |
| Psoriatic arthropathy | Certolizumab pegol | 8 | 220,912 | 95 | 211,057 | 12.43 [6.04,25.58] | $< 2.8 \times 10^{-16}$ |
| Internal haemorrhage | Rivaroxaban | 9 | 220,911 | 101 | 211,051 | 11.75 [5.94,23.23] | $< 4.2 \times 10^{-17}$ |
| Arthralgia | Certolizumab pegol | 16 | 220,904 | 175 | 210,977 | 11.45 [6.86,19.11] | $< 8.2 \times 10^{-32}$ |
| Hypoxia | Cyclophosphamide | 8 | 220,912 | 76 | 211,076 | 9.94 [4.80,20.60] | $< 3.9 \times 10^{-11}$ |
| Haemorrhagic stroke | Rivaroxaban | 11 | 220,909 | 99 | 211,053 | 9.42 [5.05,17.56] | $< 4.8 \times 10^{-15}$ |
| Rectal haemorrhage | Rivaroxaban | 22 | 220,898 | 179 | 210,973 | 8.52 [5.47,13.27] | $< 8.9 \times 10^{-29}$ |
| Cardiac arrest | Alprazolam | 15 | 220,905 | 117 | 211,035 | 8.16 [4.77,13.98] | $< 3.1 \times 10^{-17}$ |
| Neurotoxicity | Tisagenlecleucel | 14 | 220,906 | 104 | 211,048 | 7.78 [4.45,13.59] | $< 1.5 \times 10^{-14}$ |
| Hypotension | Fludarabine | 12 | 220,908 | 88 | 211,064 | 7.68 [4.20,14.03] | $< 1.5 \times 10^{-11}$ |
| Hypotension | Tisagenlecleucel | 12 | 220,908 | 87 | 211,065 | 7.59 [4.15,13.88] | $< 2.8 \times 10^{-11}$ |
| Pyrexia | Fludarabine | 30 | 220,890 | 208 | 210,944 | 7.26 [4.95,10.65] | $< 3.3 \times 10^{-31}$ |
| Neurotoxicity | Fludarabine | 16 | 220,904 | 107 | 211,045 | 7.00 [4.14,11.84] | $< 4.6 \times 10^{-14}$ |
| Tremor | Pimavanserin | 14 | 220,906 | 93 | 211,059 | 6.95 [3.96,12.20] | $< 1.5 \times 10^{-11}$ |
| Dementia | Pimavanserin | 17 | 220,903 | 103 | 211,049 | 6.34 [3.80,10.59] | $< 1.7 \times 10^{-12}$ |
| Gastrointestinal haemorrhage | Rivaroxaban | 143 | 220,777 | 855 | 210,297 | 6.28 [5.26,7.49] | $< 8.2 \times 10^{-127}$ |
| Pyrexia | Tisagenlecleucel | 34 | 220,886 | 201 | 210,951 | 6.19 [4.30,8.90] | $< 3.8 \times 10^{-27}$ |
| Confusional state | Pimavanserin | 66 | 220,854 | 367 | 210,785 | 5.83 [4.48,7.57] | $< 5.2 \times 10^{-50}$ |
| Hallucination | Pimavanserin | 210 | 220,710 | 1,145 | 210,007 | 5.73 [4.95,6.64] | $< 4.0 \times 10^{-161}$ |
| Haematuria | Rivaroxaban | 36 | 220,884 | 183 | 210,969 | 5.32 [3.72,7.61] | $< 8.2 \times 10^{-22}$ |
| Gait disturbance | Pimavanserin | 35 | 220,885 | 168 | 210,984 | 5.03 [3.49,7.23] | $< 1.1 \times 10^{-18}$ |
| Delusion | Pimavanserin | 57 | 220,863 | 269 | 210,883 | 4.94 [3.71,6.58] | $< 1.2 \times 10^{-31}$ |
| Hypotension | Cyclophosphamide | 24 | 220,896 | 113 | 211,039 | 4.93 [3.17,7.66] | $< 3.0 \times 10^{-11}$ |
| Neurotoxicity | Cyclophosphamide | 24 | 220,896 | 112 | 211,040 | 4.88 [3.14,7.59] | $< 5.1 \times 10^{-11}$ |
| Melanocytic naevus | Fingolimod | 27 | 220,893 | 118 | 211,034 | 4.57 [3.01,6.95] | $< 5.7 \times 10^{-11}$ |
| Cough | Epoprostenol | 31 | 220,889 | 134 | 211,018 | 4.52 [3.06,6.69] | $< 6.5 \times 10^{-13}$ |
| Chest pain | Epoprostenol | 41 | 220,879 | 171 | 210,981 | 4.37 [3.11,6.14] | $< 8.4 \times 10^{-17}$ |
| Arthralgia | Riociguat | 30 | 220,890 | 125 | 211,027 | 4.36 [2.93,6.50] | $< 2.2 \times 10^{-11}$ |
| Hypoacusis | Pregabalin | 33 | 220,887 | 129 | 211,023 | 4.09 [2.79,6.00] | $< 5.5 \times 10^{-11}$ |
| Cardiac arrest | Acetaminophen | 46 | 220,874 | 177 | 210,975 | 4.03 [2.91,5.57] | $< 5.2 \times 10^{-16}$ |
| Hallucination | Carbidopa | 106 | 220,814 | 404 | 210,748 | 3.99 [3.22,4.95] | $< 3.1 \times 10^{-40}$ |

| | | | | | | | |
|---|---|---|---|---|---|---|---|
| Neuropathy peripheral | Lenalidomide | 236 | 220,684 | 844 | 210,308 | 3.75 [3.25,4.34] | $< 2.5 \times 10^{-82}$ |
| Pain in jaw | Macitentan | 109 | 220,811 | 379 | 210,773 | 3.64 [2.94,4.51] | $< 5.1 \times 10^{-34}$ |
| Hallucination | Quetiapine | 47 | 220,873 | 163 | 210,989 | 3.63 [2.62,5.02] | $< 9.5 \times 10^{-13}$ |
| Pain in extremity | Epoprostenol | 55 | 220,865 | 187 | 210,965 | 3.56 [2.64,4.81] | $< 1.3 \times 10^{-14}$ |
| Chest discomfort | Macitentan | 60 | 220,860 | 201 | 210,951 | 3.51 [2.63,4.68] | $< 9.1 \times 10^{-16}$ |
| Fluid retention | Epoprostenol | 45 | 220,875 | 150 | 211,002 | 3.49 [2.50,4.87] | $< 1.0 \times 10^{-10}$ |
| Cough | Riociguat | 46 | 220,874 | 152 | 211,000 | 3.46 [2.49,4.81] | $< 1.0 \times 10^{-10}$ |
| Pyrexia | Cyclophosphamide | 79 | 220,841 | 259 | 210,893 | 3.43 [2.67,4.42] | $< 1.3 \times 10^{-20}$ |
| Urinary tract infection | Macitentan | 59 | 220,861 | 193 | 210,959 | 3.42 [2.56,4.58] | $< 1.5 \times 10^{-14}$ |
| Pain in jaw | Sildenafil | 78 | 220,842 | 255 | 210,897 | 3.42 [2.66,4.41] | $< 3.7 \times 10^{-20}$ |
| Arthralgia | Macitentan | 97 | 220,823 | 313 | 210,839 | 3.38 [2.69,4.24] | $< 3.6 \times 10^{-25}$ |
| Insomnia | Macitentan | 52 | 220,868 | 168 | 210,984 | 3.38 [2.48,4.62] | $< 5.8 \times 10^{-12}$ |
| Pain in extremity | Macitentan | 133 | 220,787 | 425 | 210,727 | 3.35 [2.76,4.07] | $< 6.6 \times 10^{-35}$ |
| Pyrexia | Macitentan | 84 | 220,836 | 257 | 210,895 | 3.20 [2.50,4.10] | $< 1.6 \times 10^{-18}$ |
| Cough | Macitentan | 176 | 220,744 | 526 | 210,626 | 3.13 [2.64,3.72] | $< 2.7 \times 10^{-40}$ |
| Chest pain | Macitentan | 126 | 220,794 | 376 | 210,776 | 3.13 [2.55,3.83] | $< 7.9 \times 10^{-28}$ |
| Neuropathy peripheral | Acyclovir | 86 | 220,834 | 251 | 210,901 | 3.06 [2.39,3.90] | $< 8.8 \times 10^{-17}$ |
| Pain in jaw | Epoprostenol | 134 | 220,786 | 378 | 210,774 | 2.95 [2.43,3.60] | $< 9.7 \times 10^{-26}$ |
| Oedema peripheral | Macitentan | 135 | 220,785 | 376 | 210,776 | 2.92 [2.40,3.55] | $< 4.3 \times 10^{-25}$ |
| Neuropathy peripheral | Dexamethasone | 134 | 220,786 | 362 | 210,790 | 2.83 [2.32,3.45] | $< 6.2 \times 10^{-23}$ |
| Dyspnoea | Epoprostenol | 152 | 220,768 | 405 | 210,747 | 2.79 [2.32,3.36] | $< 2.0 \times 10^{-25}$ |
| Neuropathy peripheral | Acetaminophen | 76 | 220,844 | 199 | 210,953 | 2.74 [2.10,3.57] | $< 2.1 \times 10^{-10}$ |
| Arthralgia | Selexipag | 93 | 220,827 | 231 | 210,921 | 2.60 [2.04,3.31] | $< 2.3 \times 10^{-11}$ |
| Hypotension | Riociguat | 103 | 220,817 | 253 | 210,899 | 2.57 [2.05,3.23] | $< 1.5 \times 10^{-12}$ |
| Neutrophilia | Clozapine | 98 | 220,822 | 239 | 210,913 | 2.55 [2.02,3.23] | $< 1.5 \times 10^{-11}$ |
| Pain in extremity | Sildenafil | 102 | 220,818 | 239 | 210,913 | 2.45 [1.95,3.09] | $< 1.4 \times 10^{-10}$ |
| Hypotension | Macitentan | 194 | 220,726 | 451 | 210,701 | 2.44 [2.06,2.88] | $< 2.0 \times 10^{-22}$ |
| Fluid retention | Macitentan | 224 | 220,696 | 515 | 210,637 | 2.41 [2.06,2.82] | $< 1.6 \times 10^{-25}$ |
| Dyspnoea | Macitentan | 672 | 220,248 | 1,518 | 209,634 | 2.37 [2.17,2.60] | $< 5.4 \times 10^{-79}$ |
| Pulmonary hypertension | Macitentan | 132 | 220,788 | 295 | 210,857 | 2.34 [1.91,2.87] | $< 1.9 \times 10^{-12}$ |
| Dyspnoea | Riociguat | 202 | 220,718 | 421 | 210,731 | 2.18 [1.85,2.58] | $< 2.9 \times 10^{-16}$ |
| Dyspnoea | Sildenafil | 375 | 220,545 | 753 | 210,399 | 2.10 [1.86,2.38] | $< 5.2 \times 10^{-29}$ |
| Dyspnoea | Selexipag | 300 | 220,620 | 577 | 210,575 | 2.02 [1.75,2.32] | $< 3.3 \times 10^{-19}$ |
| Pneumonia | Macitentan | 286 | 220,634 | 541 | 210,611 | 1.98 [1.72,2.29] | $< 5.5 \times 10^{-17}$ |
| Dyspnoea | Apixaban | 212 | 220,708 | 396 | 210,756 | 1.96 [1.66,2.31] | $< 4.5 \times 10^{-11}$ |
| Neoplasm progression | Palbociclib | 248 | 220,672 | 443 | 210,709 | 1.87 [1.60,2.19] | $< 4.9 \times 10^{-11}$ |
| Dyspnoea | MP glycol-epoetin beta | 138 | 220,782 | 32 | 211,120 | 0.24 [0.17,0.36] | $< 3.6 \times 10^{-11}$ |
| Pyrexia | Deflazacort | 50 | 220,870 | 0 | 211,152 | 0.00 [0.00,0.00] | $< 1.8 \times 10^{-10}$ |
| Haematotoxicity | Carboplatin | 0 | 220,920 | 46 | 211,106 | - - | $< 3.0 \times 10^{-10}$ |
| Cardiac arrest | Remdesivir | 0 | 220,920 | 128 | 211,024 | - - | $< 9.5 \times 10^{-36}$ |
| Haematotoxicity | Cisplatin | 0 | 220,920 | 53 | 211,099 | - - | $< 2.0 \times 10^{-12}$ |
| Insomnia | Elexacaftor | 0 | 220,920 | 55 | 211,097 | - - | $< 4.8 \times 10^{-13}$ |
| Hypotension | Remdesivir | 0 | 220,920 | 117 | 211,035 | - - | $< 2.5 \times 10^{-32}$ |
| Respiratory failure | Remdesivir | 0 | 220,920 | 119 | 211,033 | - - | $< 6.0 \times 10^{-33}$ |
| Type I hypersensitivity | Cetirizine | 0 | 220,920 | 56 | 211,096 | - - | $< 2.3 \times 10^{-13}$ |
| Bradycardia | Remdesivir | 0 | 220,920 | 125 | 211,027 | - - | $< 8.1 \times 10^{-35}$ |
| Insomnia | Istradefylline | 0 | 220,920 | 49 | 211,103 | - - | $< 3.5 \times 10^{-11}$ |
| Hypoxia | Remdesivir | 0 | 220,920 | 100 | 211,052 | - - | $< 4.9 \times 10^{-27}$ |
| Type I hypersensitivity | Famotidine | 0 | 220,920 | 55 | 211,097 | - - | $< 4.8 \times 10^{-13}$ |
| Dyspnoea | Remdesivir | 0 | 220,920 | 67 | 211,085 | - - | $< 9.0 \times 10^{-17}$ |
| Shock haemorrhagic | Rivaroxaban | 0 | 220,920 | 49 | 211,103 | - - | $< 3.5 \times 10^{-11}$ |
| Tremor | Istradefylline | 0 | 220,920 | 47 | 211,105 | - - | $< 1.4 \times 10^{-10}$ |
| Type I hypersensitivity | Oxaliplatin | 0 | 220,920 | 70 | 211,082 | - - | $< 1.0 \times 10^{-17}$ |
| Hallucination | Istradefylline | 0 | 220,920 | 78 | 211,074 | - - | $< 3.4 \times 10^{-20}$ |
| Haematotoxicity | Thiotepa | 0 | 220,920 | 46 | 211,106 | - - | $< 3.0 \times 10^{-10}$ |
